# Extended Effects of a Wearable Sensory Prosthesis on Gait, Balance Function and Falls After 26 Weeks of Use in Persons with Peripheral Neuropathy and High Fall Risk – The walk2Wellness Trial

**DOI:** 10.1101/2022.04.28.22274328

**Authors:** Lars IE Oddsson, Teresa Bisson, Helen S Cohen, Ikechukwu Iloputaife, Laura Jacobs, Doris Kung, Lewis A Lipsitz, Brad Manor, Patricia McCracken, Yvonne Rumsey, Diane M Wrisley, Sara R Koehler-McNicholas

## Abstract

**Background:** We recently reported that individuals with impaired plantar sensation and high fall risk due to sensory peripheral neuropathy (PN) improved gait and balance function following 10 weeks of use of Walkasins^®^, a wearable lower limb sensory prosthesis that provides directional tactile stimuli related to plantar pressure measurements during standing and walking (RxFunction Inc., MN, USA). Here, we report 26-week outcomes and compare pre- and in-study fall rates. We expected improvements in outcomes and reduced fall rates reported after 10 weeks of use to be sustained.

**Methods:** Participants had clinically diagnosed PN with impaired plantar sensation, high fall risk (Functional Gait Assessment, FGA score <23) and ability to sense tactile stimuli above the ankle at the location of the device. Additional outcomes included 10m Gait Speed, Timed Up&Go (TUG), Four-Stage Balance Test, and self-reported outcomes, including Activities-Specific Balance Confidence scale and Vestibular Disorders Activities of Daily Living Scale. Participants tracked falls using a calendar.

**Results:** We assessed falls and self-reported outcomes from 44 individuals after 26 weeks of device use; 30 of them conducted in-person testing of clinical outcomes. Overall, improvements in clinical outcomes seen at 10 weeks of use remained sustained at 26 weeks with statistically significant increases compared to baseline seen in FGA scores (from 15.0 to 19.2), self-selected gait speed (from 0.89 m/s to 0.97 m/s), and 4-Stage Balance Test (from 25.6s to 28.4s), indicating a decrease in fall risk. Non-significant improvements were observed in TUG and fast gait speed. Overall, 39 falls were reported; 31 of them did not require medical treatment and four caused severe injury. Participants who reported falls over 6 months prior to the study had a 43% decrease in fall rate during the study as compared to self-report 6-month pre-study (11.8 vs. 6.7 falls/1000 patient days, respectively, p<0.004), similar to the 46% decrease reported after 10 weeks of use.

**Conclusion:** A wearable sensory prosthesis can improve outcomes of gait and balance function and substantially decreases incidence of falls during long-term use. The sustained long-term benefits in clinical outcomes reported here lessen the likelihood that improvements are placebo effects.

**Trial registration:** ClinicalTrials.gov (#NCT03538756)

## 1 Introduction

Persistent problems with gait and balance function may lead to falls, fractures, and other serious injuries in older adults (Bergen, Stevens et al. 2016, Ganz and Latham 2020, Moreland, Kakara et al. 2020). In fact, clinical observations on the problem of falls and fall prevention go back to at least 1948 when Sheldon (Sheldon 1948) presented data in *The Lancet* on “Liability to Falls” in older adults, concluding that falling was “a clinical problem which has never received the attention it deserves.” Today, we appreciate that causes of falls are complex and require a multifactorial clinical approach to arrive at a correct diagnosis and to provide specific and relevant therapeutic interventions (Cohen, Miller et al. 2015).

To address the problem of falls in older adults, various clinical practice guidelines for falls prevention and management have been developed (Montero-Odasso, Kamkar et al. 2021). The Centers for Disease Control and Prevention (CDC), for example, has established “STEADI” (STEADI 2021), an evidence-based initiative to address fall prevention based on a curation of decades of research activities (Stevens and Phelan 2013). At this time, advice provided through the STEADI initiative includes “Talk to your doctor”; “Do Strength and Balance Exercises”; “Have Your Eyes Checked”; and “Make Your Home Safer”(CDC 2021). At a minimum, health care providers should screen their patients for fall risk, including older adults and individuals with neurodegenerative diseases that affect balance function. Simple interventions that should be considered to help reduce the risk for falls include medication management and vitamin D supplements to improve bone, muscle, and nerve health (Bergen, Stevens et al. 2016). Further treatment of gait and balance issues may involve use of canes, walkers, and rehabilitation interventions with balance exercises (Richardson, Sandman et al. 2001, Halvarsson, Franzén et al. 2013, Melzer and Oddsson 2013, Ganz and Latham 2020), including Tai-Chi (Li and Manor 2010, Manor, Lough et al. 2014, Quigley, Bulat et al. 2014). Any functional benefits related to fall rates and gait function after long-term use of bilateral ankle foot orthoses appear limited (Wang, Goel et al. 2019).

In addition to other age-related risk factors, sensory peripheral neuropathy (PN), leading to impaired plantar sensation, is an independent risk factor for falls (Richardson and Hurvitz 1995, Koski, Luukinen et al. 1998, Reeves, Orlando et al. 2021), irrespective of whether the underlying cause is due to diabetes (Mustapa, Justine et al. 2016, Vinik, Camacho et al. 2017), chemotherapy (Winters-Stone, Horak et al. 2017), or an unknown factor (Riskowski, Quach et al. 2012). The prevalence of PN in the US population for those over age 40 is nearly 15% (Gregg, Sorlie et al. 2004) and 28.4% in persons with diabetes (Hicks, Wang et al. 2021). The incidence of injuries due to falls is 15 times higher in patients with diabetic PN than in healthy individuals (Cavanagh, Derr et al. 1992). Furthermore, polyneuropathy contributes independently to functional impairments, including difficulty walking and a tendency to fall (Hoffman, Staff et al. 2015), and those individuals more often incur fall-related injuries (Hanewinckel, Drenthen et al. 2017). A prospective study of older individuals with PN found that 65% fell during a one-year period and 30% reported an injury from a fall (DeMott, Richardson et al. 2007).

Mechanoreceptors play an important role in proprioception (Fallon, Bent et al. 2005, Vaughan 2021) and loss of sensory information from the lower limbs, including plantar mechanoreceptors alters balance performance (Meyer, Oddsson et al. 2004, Meyer, Oddsson et al. 2004). Furthermore, this sensory impairment affects the automaticity of walking, thereby increasing the need for cognitive attention to gait and mobility activities (Paul, Ada et al. 2005, Clark, Christou et al. 2014, Clark 2015). Furthermore, several studies have reported problems with gait and balance function and the increased risk of falls in patients with PN (Menz, Lord et al. 2004, DeMott, Richardson et al. 2007, Dixon, Knight et al. 2017, Lipsitz, Manor et al. 2018).

Additionally, low gait speed is a risk factor for falls (Studenski, Perera et al. 2003, Montero-Odasso, Schapira et al. 2005), is an indicator of frailty (Kim, Glynn et al. 2019), and is even a predictor of hospitalization and disability (Cesari, Kritchevsky et al. 2005, Dumurgier, Elbaz et al. 2009) as well as survival (Studenski, Perera et al. 2011). In fact, interventions designed to improve gait speed may actually increase survival (Hardy, Perera et al. 2007). Older adults with sensory impairment related to PN show a decline in gait speed that is four times higher than in healthy aging (Buracchio, Dodge et al. 2010, Lipsitz, Manor et al. 2018).

Although strength and balance training in patients with PN may help reduce fall risk falls (Ites, Anderson et al. 2011, Tofthagen, Visovsky et al. 2012, Streckmann, Zopf et al. 2014, Morrison, Simmons et al. 2018), strength training in patients with PN appears to have less impact on balance (Streckmann, Zopf et al. 2014). Its effects are mainly compensatory and do not address impaired somatosensation, the root cause of balance problems related to PN. Furthermore, balance training activities must be specific (Oddsson, Boissy et al. 2007) and must be conducted with sufficient intensity and frequency (Lipsitz, Macklin et al. 2019) to be helpful, or benefits will be limited or absent (Kruse, Lemaster et al. 2010, Lipsitz, Manor et al. 2018, Lipsitz, Macklin et al. 2019). Clear guidelines regarding frequency of balance exercises are currently lacking although three sessions a week may be a minimum necessary to see an improvement (Kruse, Lemaster et al. 2010). Thus, although “Falls can be prevented” (CDC 2021), they continue to be a large problem in older adults, indicating a continued need for novel solutions.

We recently reported from our multi-site clinical trial (walk2Wellness, NCT #03538756, www.clinicaltrials.gov) that individuals with PN and a high risk of falls improved their Functional Gait Assessment (FGA) scores, (Wrisley, Marchetti et al. 2004), Gait Speed, and Timed Up & Go times following 10 weeks of home-based use of a wearable sensory prosthesis intended to substitute nerve function related to impaired plantar sensation (Figure 1, Walkasins by RxFunction Inc., Eden Prairie, MN, USA), (Oddsson, Bisson et al. 2020). Previously, we found similar meaningful improvements in FGA scores and gait speed in participants with PN in a randomized crossover trial conducted in-clinic (Koehler-McNicholas, Danzl et al. 2019), findings that were recently referred to as “presumably emulating” signals by fast-adapting cutaneous afferents (Vaughan 2021). We also reported a decrease in the number of fall risk factors as well as fall rate from baseline to 10 weeks for individuals who reported falls in the six months preceding study participation (Oddsson, Bisson et al. 2020).

**Figure 1.**
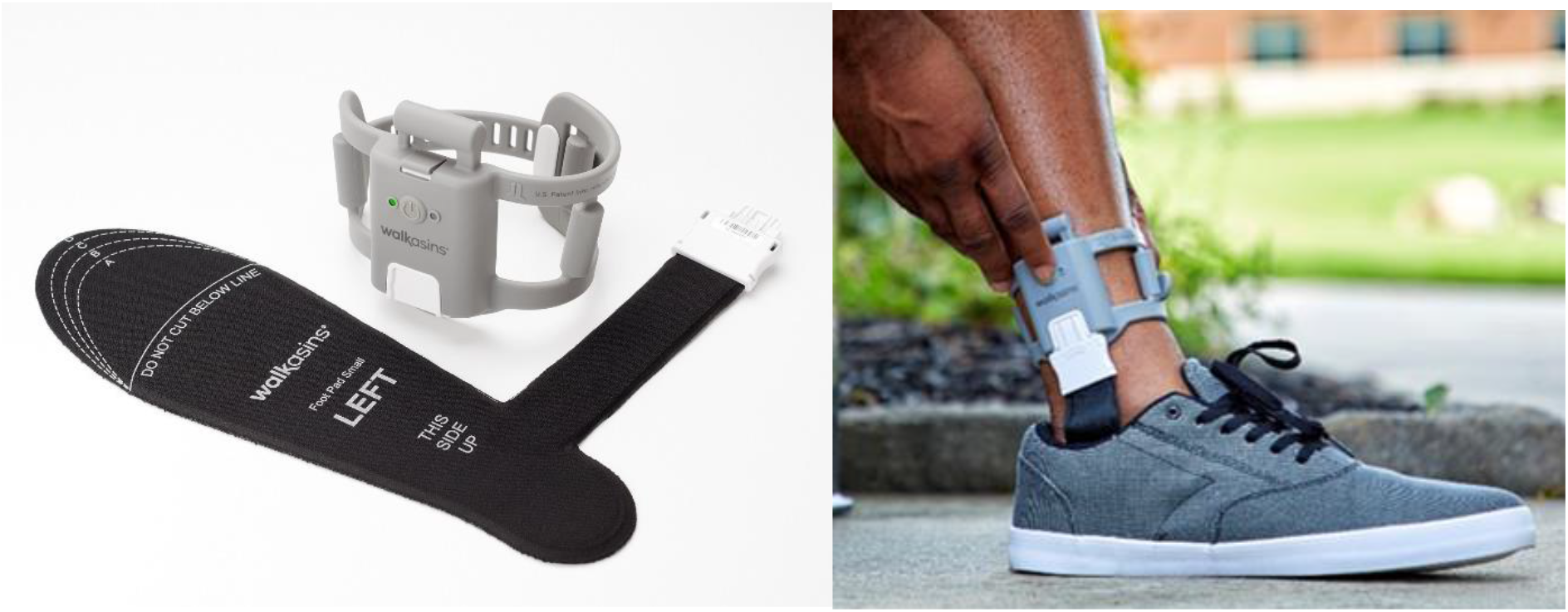
(Left) The two components of the Walkasins device, the pressure sensitive flexible foot pad that is placed in the shoe and connects to the leg unit that contains a rechargeable battery, a microprocessor, supporting electronics, and four tactile stimulators. The embedded software algorithm evaluates pressure data and activates tactile stimulators at relevant times during standing and walking to signal balance-related information to the afferent nervous system. (Right) A Walkasins user wearing the device in the process of turning it on. The Walkasins system is worn bilaterally (unilateral components depicted).

Here we report extended long-term use data from the walk2Wellness trial for 44 participants after 26 weeks of Walkasins use. Furthermore, we analyzed falls during the 26 weeks of device use and compared to patient self-reports of falls from the six months preceding participation in the study. We expected improvements in clinical outcomes and lower fall rate seen after 10 weeks (Oddsson, Bisson et al. 2020) to be sustained at 26 weeks.

## 2 Materials and Methods

Detailed descriptions of materials and methods have been reported previously (Oddsson, Bisson et al. 2020), and are summarized below. The measures we use have been validated (Oddsson, Bisson et al. 2020) and our primary outcome measure, the FGA, is recommended for assessment of walking balance in adults with neurologic conditions by current physical therapy Clinical Practice Guidelines (Moore, Potter et al. 2018). The walk2Wellness study is registered at ClinicalTrials.gov (#NCT03538756). Figure 1 shows a picture of the Walkasins device components with a brief description of its functionality.

### 2.1 Participants

The walk2Wellness clinical trial received approval for human subject research from Advarra IRB (formerly Quorum Review) for four sites and from the IRB Subcommittee, the Subcommittee on Research Safety, and the Research and Development Committee of the Minneapolis VA Health Care System (MVAHCS). Recruitment for the walk2Wellness trial began in October 2018 and ended at the last site in April 2021. Because of COVID-19 disruptions, the study ended earlier than planned at four sites; the fifth site completed the trial but with a reduced enrollment goal. Study site team members posted fliers at community businesses and clinics to advertise the trial; in addition, some of the sites received referrals from physicians who were informed about the study.

To participate in the trial, individuals met the following inclusion criteria: male or female; ages 21-90; a formal diagnosis of sensory PN prior to participating in the study as indicated by their medical record or a letter by a physician; self-reported problems with balance; ability for transfers or ambulation on level surfaces at fixed cadence as assessed by trained study personnel; FGA score <23, the cut-off score for high fall risk (Wrisley and Kumar 2010); ability to understand and provide informed consent; foot size to allow the Walkasins device to function properly, and ability to complete all functional outcome measures without the use of an assistive device in order to ensure sufficient motor function. Participants could use an assistive device at their discretion during their daily activities.

Individuals were excluded if they were unable to perceive the tactile stimuli from the Walkasins Leg Unit (Figure 1) or if they used an ankle-foot orthosis for ambulation that prevented donning the device. In addition, individuals having any of the following conditions were excluded from participation: acute thrombophlebitis; deep vein thrombosis; untreated lymphedema; a lesion of any kind, swelling, infection, inflamed area of skin, or eruptions on the lower leg near placement of the device; foot or ankle fractures; severe peripheral vascular disease; or any musculoskeletal or neurological condition that would prohibit use of the device, as determined by a clinician. Because of a potential to overload the pressure sensors in the foot pad, individuals weighing over 136 kg (300 lbs) were excluded from participation. The criteria to participate in the study were similar to receiving a prescription for the device in a clinic. Potential participants also agreed not to initiate any balance training (e.g., Tai-Chi, etc.) or balance-related therapy during the first 10 weeks of the trial (Oddsson, Bisson et al. 2020). Furthermore, participants were blinded to their outcomes during the trial and not shared by study personnel.

### 2.2 Study Procedures and Outcome Measures

Before undergoing any study-related procedures, individuals signed the IRB-approved consent form and HIPAA authorization. After enrollment, site staff tested whether participants were able to perceive the tactile stimuli from the Walkasins Leg Unit. Anyone unable to feel the tactile stimuli was excluded from continued participation in the study. Individuals who were able to feel the tactile stimuli from the Walkasins Leg Unit bilaterally then completed a medical history form to assess common health issues and systemic diseases. They also provided information on falls they had experienced in the previous six and twelve months, including the number of falls and any injuries they sustained as a result. In addition, participants provided a list of their medications (medication name, indication, dose, and frequency), which was updated over the course of the study.

Participants then completed the Activities-Specific Balance Confidence (ABC) Questionnaire (Powell and Myers 1995) to assess levels of balance self-confidence and the Vestibular Activities of Daily Living Scale (VADL), (Cohen, Kimball et al. 2000) to evaluate the effects of vertigo and balance disorders on their ability to perform everyday activities independently. At baseline, week 10, and week 26, site personnel also assessed participants using the Weinstein Enhanced Sensory Test (WEST) monofilament foot test (0.5g, 2g, 10g, 50g, and 200g), as described previously (Oddsson, Bisson et al. 2020). Staff also performed a vibration sensation test using a Rydel-Seiffer Tuning Fork to document loss of sensation (Kästenbauer, Sauseng et al. 2004, Oddsson, Bisson et al. 2020). Scoring values ≤4 at the first metatarsal joint is categorized as abnormal (Kästenbauer, Sauseng et al. 2004).

During the baseline visit, participants donned the Walkasins devices and performed a standardized set of standing and walking balance activities focused on orientation and familiarization with the device, referred to as the Walkasins Learning Protocol (Koehler-McNicholas, Danzl et al. 2019, Oddsson, Bisson et al. 2020). Trained site personnel then assessed participants on the FGA, 10-Meter Walk Test (Perera, Mody et al. 2006), Timed Up & Go (TUG) (Mathias, Nayak et al. 1986), and 4-Stage Balance Test (CDC 2021). The clinical outcomes were standardized and performed by study personnel who were trained by one of the investigators (DW). A sponsor representative conducted observation visits periodically during the study to ensure standardization across the sites (LJ).

After finishing the clinical outcome tests, participants completed the following self-reported outcome measures to provide information about their quality of life, pain level, and engagement in social activities: Patient Health Questionnaire (PHQ-9) (Kroenke, Spitzer et al. 2001), PROMIS Pain Interference Short Form 6b (Askew, Cook et al. 2016), PROMIS Pain Intensity Form 1a, Ability to Participate Short Form 8a (Hahn, Kallen et al. 2016), and Satisfaction with Participation in Social Roles Short Form 8a (Hahn, DeWalt et al. 2014, Hahn, Beaumont et al. 2016). At the conclusion of the baseline visit, site staff members trained the participants on caring for the device.

Participants left the baseline visit with the device and a calendar on which to note any fall events they experienced as well as their use of Walkasins. For consistency across sites, we used the World Health Organization’s definition of a “fall”: “A fall is an event which results in a person coming to rest inadvertently on the ground or floor or other lower level.” When participants reported falls, study staff recorded the following: the fall date; severity (no injury, minor injury, or major injury); the level of treatment the individual sought or received, if any; the cause or causes of the fall (environmental factor, lost balance, lost consciousness, lost strength/felt weak, slipped, tripped, on new medication, other, unknown); and a description of the fall.

Unless prevented from doing so because of COVID-19 site restrictions, participants returned for in-person visits at weeks 2, 6, 10, and 26. These visits followed most of the same procedures as the baseline visit: balance questionnaires, clinical assessments, and self-reported outcome measures. As we have previously reported (Oddsson, Bisson et al. 2020) we calculated ABC to FGA ratios at each assessment to measure the degree of internal self-perception of balance capability (ABC score) in relation to the externally observed walking balance performance (FGA score). We noted that a ratio of around 3.3 would be “expected” based on the maximum scores of the two scales. This construct aligns with a multifactorial causation model for falls emphasizing the importance of a “Realistic Appraisal of One’s Own Abilities”, (Hadjistavropoulos, Delbaere et al. 2011).

Between the in-person visits at weeks 10 and 26, study sites contacted participants via telephone at weeks 14, 18, and 22 to remind them of study requirements and to collect follow-up information regarding health changes, falls, adverse events, pain scores, device usage, and device functioning, as well as whether they had begun any physical therapy for their balance problems. If participants reported adverse events and/or falls during these contacts, site personnel recorded the details on the appropriate case report forms in REDCap Cloud, the electronic data capture system used in the study (https://www.redcapcloud.com/, Encinitas, CA, United States).

### 2.1 Statistical Analysis and Availability of Data

An analysis of outcomes after 2, 6, and 10 weeks, with 10 weeks being the primary endpoint, was presented earlier (Oddsson, Bisson et al. 2020). Here we focus on outcomes after extended long-term device use and compare outcomes to baseline for individuals participating in 26 weeks of device use. Graphical data from 2, 6 and 10 weeks are provided below for illustration. We calculated descriptive statistics (mean and standard deviation or median as appropriate) for all variables and tested for normality with the Shapiro-Wilk’s test. We applied the two-proportion Z-test to compare proportion-based measures. We conducted a post-hoc analysis to compare participants at baseline who reported falls in the previous six months (Pre-Fallers, n=25) to those who did not (Pre-NonFallers, n=19). Their baseline characteristics were compared using a t-test for independent samples or a Mann-Whitney U test if data was not normally distributed based on a Shapiro-Wilk’s test.

To compare baseline variables to those after 26 weeks of device use, we conducted a t-test for dependent samples or Wilcoxon signed-rank test in case variables were not normally distributed. We applied a Bonferroni’s adjustment of significance levels for correlated measures, ranging from p<0.0167 (0.05/3 for three comparisons) for a full correction (non-correlated measures, r=0) and p<0.05 for perfectly correlated measures (r=1) (Uitenbroek 1997). We calculated effect sizes using Cohen’s drm according to recommendations by Lakens (Lakens 2013) and interpreted them according to Cohen (Cohen 1988), i.e. 0.2 represents a small effect, 0.5 a medium effect, and 0.8 a large effect size. Ninety-five percent confidence intervals of effect sizes were estimated according to Algina (Algina, J et al. 2005).

We performed statistical analysis with the Analysis-ToolPak module in Microsoft Excel 2016 and the Real Statistics Resource Pack software, release 6.8 (Zaiontz 2020). Baseline characteristics of participants in the study from the four clinical sites are presented in Table 1. We have pooled the data for the continued analysis presented here. The raw data supporting the conclusions of this article will be made available to qualified researchers without undue reservation.

**Table 1.** Characteristics of the 44 individuals from the four different clinical sites enrolled in the study who reached 26 weeks of participation. Values represent Mean (Standard Deviation). #ChrD - Number of Chronic Diseases.

|  | n | Age<br>(yrs) | Height<br>(m) | Weight<br>(kg) | #ChrD | FGA<br>Score | Gait Speed<br>Self-selected<br>(m/s) | Gait Speed<br>Fast (m/s) | TUG<br>(s) | 4-Stage<br>Balance<br>Test (s) | ABC<br>Score |
| --- | --- | --- | --- | --- | --- | --- | --- | --- | --- | --- | --- |
| **VAMC** | 18 | 74.4<br>(6.0) | 1.76<br>(0.07) | 96.9<br>(14.1) | 9.6<br>(2.3) | 15.4<br>(4.1) | 0.82<br>(0.13) | 1.27<br>(0.36) | 13.5<br>(2.9) | 24.8<br>(7.4) | 68.2<br>(15.8) |
| **Baylor** | 11 | 74.6<br>(9.4) | 1.75<br>(0.08) | 87.0<br>(15.2) | 8.2<br>(2.9) | 15.4<br>(4.5) | 0.88<br>(0.25) | 1.33<br>(0.39) | 13.3<br>(6.3) | 28.1<br>(8.0) | 55.6<br>(18.8) |
| **M Health**<br>**Fairview** | 5 | 71.2<br>(10.7) | 1.75<br>(0.14) | 84.4<br>(12.0) | 6.8<br>(5.3) | 15.6<br>(2.1) | 1.07<br>(0.21) | 1.32<br>(0.20) | 10.5<br>(1.4) | 26.4<br>(3.8) | 64.1<br>(7.0) |
| **Harvard** | 10 | 71.5<br>(8.0) | 1.76<br>(0.13) | 86.7<br>(16.9) | 6.6<br>(3.5) | 15.0<br>(3.0) | 0.95<br>(0.31) | 1.25<br>(0.45) | 12.8<br>(3.3) | 25.3<br>(7.6) | 60.8<br>(15.3) |

## 3 Results

### 3.1 Enrollment and Allocation

Our previous report with 45 participants at the 10 week primary endpoint (Oddsson, Bisson et al. 2020) was published early due to the unknown circumstances at the time related to COVID-19, which had caused recruitment holds and suspension of in-person testing activities at the research sites. Over time, the trial continued with the enrollment of additional participants who are part of the current report as illustrated in the flow chart in Figure 2. Figure 2 shows the flow chart for the study as it was continued from the 10-week primary endpoint assessment that we have previously reported (Oddsson, Bisson et al. 2020). Text boxes with dashed lines in Figure 2 indicate enrollment numbers for the initial 10-week report.

**Figure 2.**
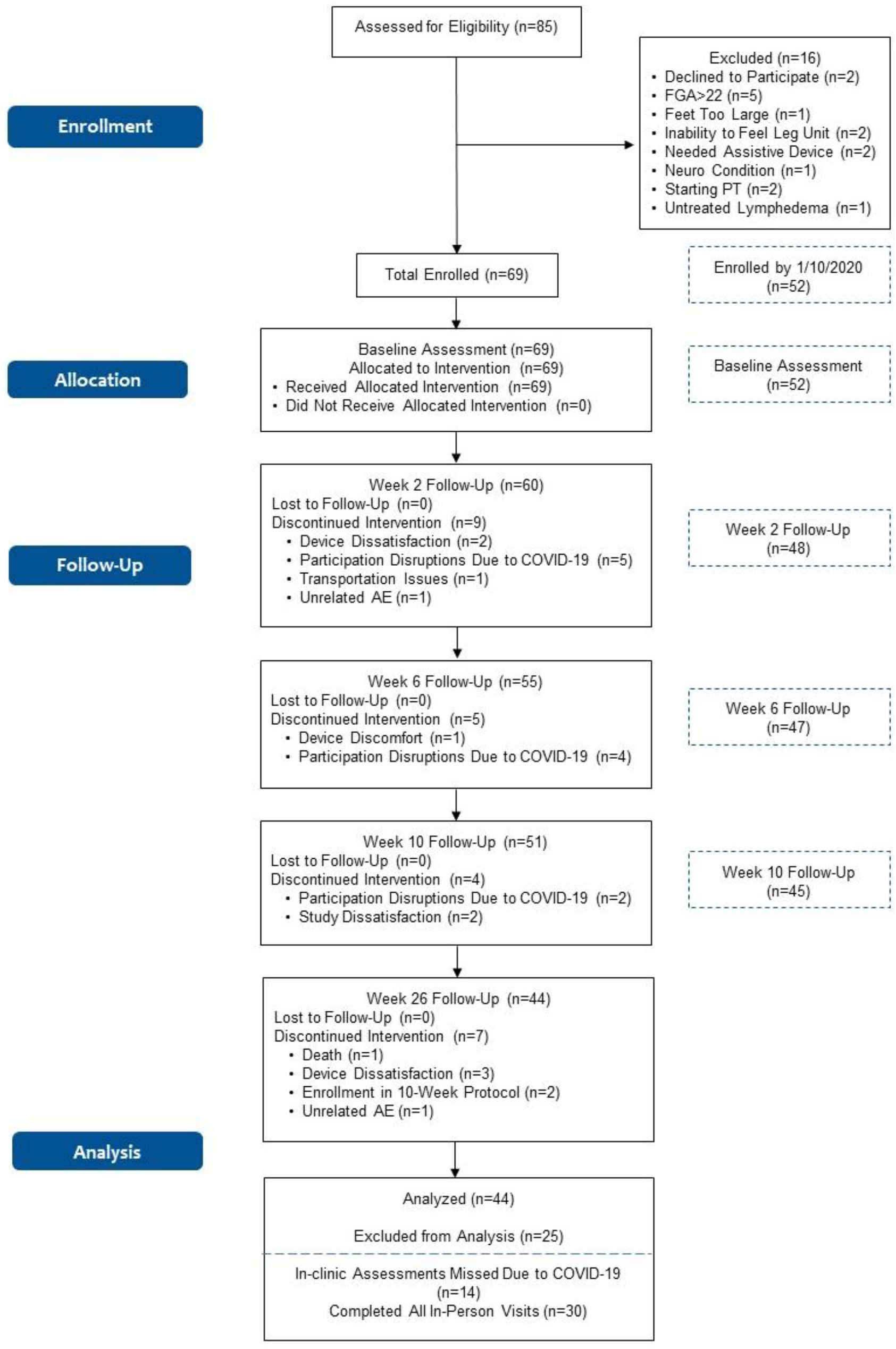
Flowchart of the walk2Wellness trial. Data for the primary endpoint at 10 weeks, shown to the right for reference, were published earlier (Oddsson, Bisson et al. 2020). Due to COVID-19 site lockdowns, in-person outcomes could be assessed from 30 of the 44 participants. Self-reported outcomes and fall events were assessed from all 44 participants who reached the 26-week follow-up visit.

Of the 85 individuals who were assessed for eligibility, a total of 69 participants were enrolled in the trial (17 more than in previous report, cf. Figure 2). Sixty participants took part in the 2-week follow up visit, 55 in the 6-week follow-up visit, and 51 in the 10-week follow-up visit (six more than in the previous report). Eleven of the 18 participants who discontinued interventions through the 10-week follow-up visit stopped participation due to circumstances related to COVID-19 (cf. Figure 2). (Note: due to the timing of study initiation at Johns Hopkins and the onset of the pandemic, the three participants enrolled at the site were unable to complete most in-person assessments. When the situation did not improve, the sponsor decided to terminate Johns Hopkins as a research site. None of the participants had completed follow-up datasets through week 10, so their results were not included in this analysis.)

Forty-four of the 51 participants who reached the 10-week follow-up assessment were analyzed after their 26-week assessment. Due to circumstances related to COVID-19, including lockdowns at the research sites, 14 of the 44 participants were unable to participate in the 26-week in-person outcomes testing (FGA, 10MWT, TUG, and 4-Stage Balance Test). All 44 participants, however, were assessed on self-reported outcomes over the phone, and all 44 participants provided reports of falls throughout the 26-week period. Consequently, we report baseline assessment of clinical outcomes for all 44 participants, comparing those who reported falling in the prior six months (Pre-Fallers) to those who did not (Pre-NonFallers). We also present data over time for the 30 participants who completed the 26-week in-person assessment (weeks 0, 2, 6, 10 and 26, Figure 2), comparing the baseline assessments of those 30 participants to their 26-week assessment.

Seven of the 44 participants discontinued the intervention after the 10-week assessment (cf. Figure 2); one passed away; two had been enrolled in solely the 10-week protocol; three expressed device dissatisfaction, and one experienced an unrelated adverse event.

### 3.2 Baseline Characteristics and Outcomes

Baseline characteristics for the 44 individuals who were evaluated at 26 weeks are shown in Table 1 and were substantially similar to those reported previously (Oddsson, Bisson et al. 2020). Participants across all sites were pooled for the continued analysis.

The baseline characteristics of the 44 participants are shown in Table 2. Characteristics are also reported separately for participants who reported falling in the six months preceding the study (Pre-F, n=25) and participants who did not report a fall (Pre-NF, n=19). Overall, characteristics reported previously for participants entering the trial were maintained (Oddsson, Bisson et al. 2020) except for some substantiated differences between the Pre-F and Pre-NF participants that were now statistically significant (Table 2). In addition to previous observations, baseline self-selected gait speed and ABC scores were significantly different between the Pre-F and Pre-NF groups, 0.83 m/s vs. 0.97 m/s and 57.8% vs. 69.6%, respectively (Table 2). A higher observed mean value in PHQ-9 score for the Pre-F group was nearly statistically significant (5.7 vs. 3.3, respectively, p=0.052). Mean values for all PROMIS measures were near 50, and any differences were well within 10 (Table 2) corresponding to one standard deviation for these measures, indicating any observed differences were minor (Askew, Cook et al. 2016, Hahn, Beaumont et al. 2016, Hahn, Kallen et al. 2016).

**Table 2.** Baseline characteristics for participants reaching the 26 Week assessment (n=44). Participants who reported having fallen in the past six months (Pre-F) and those who did not (Pre-NF) are reported separately. Values represent Mean (Standard Deviation). Column p-level shows significance level for comparison between the Pre-F and Pre-NF groups. In bold if p<0.05. *Fall-risk factors assessed in the current study included, recent history of falls (Tinetti and Kumar 2010), PN diagnosis (Richardson and Hurvitz 1995), FGA score<23 (Wrisley and Kumar 2010), TUG >12 s (CDC 2017), 4-Stage Balance Test<30s (CDC 2017), Gait Speed <0.7m/s (Studenski, Perera et al. 2003, Montero-Odasso, Schapira et al. 2005), and ABC score <67% (Lajoie and Gallagher 2004).

| Baseline Assessment | All<br>n=44 | Pre-F<br>n=25 | Pre-NF<br>n=19 | p-level |
| --- | --- | --- | --- | --- |
| Gender Female (n) | 8 of 44 (18%) | 8 of 25 (32%) | 0 of 19 (0%) | **<0.01** |
| Use of Assistive Device (n) | 25 of 44 (57%) | 18 of 25 (72%) | 7 of 19 (37%) | **0.02** |
| Gait Speed Self-selected $< 0.7$ m/s (n) | 8 of 44 (18%) | 7 of 25 (28%) | 1 of 19 (5%) | 0.052 |
| Timed Up and Go $> 12$ s (n) | 21 of 44 (48%) | 16 of 25 (64%) | 5 of 19 (26%) | **0.013** |
| 4-Stage Balance Test $< 30$ s (n) | 28 of 44 (64%) | 18 of 25 (72%) | 10 of 19 (53%) | 0.19 |
| ABC Score $< 67\%$ (n) | 24 of 44 (54%) | 17 of 25 (68%) | 7 of 19 (37%) | **0.04** |
| Fallen in Last 6 Months (n) | 25 of 44 (57%) | 25 of 25 (100%) | 0 of 19 (0%) | n/a |
| Fallen in Last 12 Months (n) | 31 of 44 (70%) | 25 of 25 (100%) | 6 of 19 (32%) | **<0.0001** |
| Number of Falls 6 Months | 53 | 53 | 0 | n/a |
| Number of Falls 12 Months | 106 | 97 of 106 (92%) | 9 of 106 (8%) | **<0.0001** |
|  | Mean (SD)<br>n=44 | Mean (SD)<br>n=25 | Mean (SD)<br>n=19 | p-level |
| Age (yrs) | 73.5 (7.8) | 73.1 (7.9) | 73.9 (7.8) | 0.72 |
| Height (m) | 1.76 (0.09) | 1.76 (0.12) | 1.76 (0.05) | 0.96 |
| Weight (kg) | 90.7 (15.3) | 89.7 (17.3) | 92.0 (12.5) | 0.63 |
| BMI (kg/m<sup>2</sup>) | 29.4 (4.6) | 29.0 (5.2) | 29.8 (3.8) | 0.59 |
| FGA Score | 15.3 (3.7) | 13.9 (3.4) | 17.3 (3.3) | **<0.002** |
| Gait Speed, Self-selected (m/s) | 0.89 (0.23) | 0.83 (0.23) | 0.97 (0.21) | **<0.05** |
| Gait Speed, Fast (m/s) | 1.29 (0.36) | 1.13 (0.29) | 1.50 (0.35) | **<0.0005** |
| TUG (s) | 12.9 (4.0) | 14.1 (4.4) | 11.4 (2.8) | **<0.026** |
| 4-Stage Balance Test (s) | 25.9 (7.2) | 24.5 (6.3) | 27.8 (8.1) | 0.14 |
| Fall-Risk Factors* (n of 7) | 3.4 (1.5) | 4.3 (0.9) | 2.2 (1.1) | **<0.0001** |
| # Chronic Conditions | 8.3 (3.3) | 8.8 (3.3) | 7.5 (3.3) | 0.21 |
| ABC Score (%) | 62.9 (16.2) | 57.8 (14.0) | 69.6 (16.8) | **<0.02** |
| VADL Mean Score | 3.7 (1.10) | 4.1 (1.0) | 3.1 (0.8) | **<0.001** |
| VAS Pain Score (0-10) | 2.9 (2.1) | 3.0 (2.1) | 2.8 (2.2) | 0.89 |
| PHQ-9 | 4.7 (4.0) | 5.7 (4.5) | 3.3 (2.9) | 0.052 |
| Pain Interference<br>PROMIS® 6b | 50.9 (8.1) | 52.7 (7.7) | 48.5 (8.1) | 0.09 |
| Satisfaction Social Roles<br>PROMIS® 8a | 49.8 (7.0) | 49.8 (6.7) | 49.9 (7.5) | 0.96 |
| Ability to Participate<br>PROMIS® 8a | 49.9 (7.4) | 48.4 (6.7) | 51.8 (8.0) | 0.13 |

### 3.3 Clinical Outcomes

Improvements in FGA scores were seen across all individuals, irrespective of their baseline FGA scores and appeared similar after 2-, 6-, 10- and 26-weeks of device use (Figure 3). This finding is indicated by the regression lines in Figure 3 being shifted above the line of equality. Figure 3 also illustrates similar improvements in FGA scores for the Pre-F group and the Pre-NF group (filled vs. open symbols). Interestingly, the slope of the regression line was <1 at all assessments indicating larger improvements in the lower range of baseline FGA scores mostly representing Pre-F group participants (cf. Table 3 and further below). After 26 weeks of device use, FGA scores increased across all individuals from 15.0 to 19.2 (p<0.00001, Table 3) indicating a large effect size (Cohen’s drm=1.38, Table 3). Furthermore, self-selected gait speed increased from 0.89 m/s to 0.97 m/s (p=0.02), a medium effect size (Cohen’s drm=0.49); and the 4-Stage Balance Test improved from 25.6 s to 28.4 s (p<0.01), representing a small effect size (Cohen’s drm=0.32). An increase seen in fast gait speed (1.30 m/s to 1.37 m/s) did not reach statistical significance (p=0.07). Changes in Rydel-Seiffer tuning fork testing scores suggested a small decrease in sensitivity at the site of the lateral malleolus (mean 3.8 to 3.2, p=0.032, Cohen’s drm=0.32), while a noted decrease at the MTP joint did not reach statistical significance (p=0.07).

**Figure 3.**
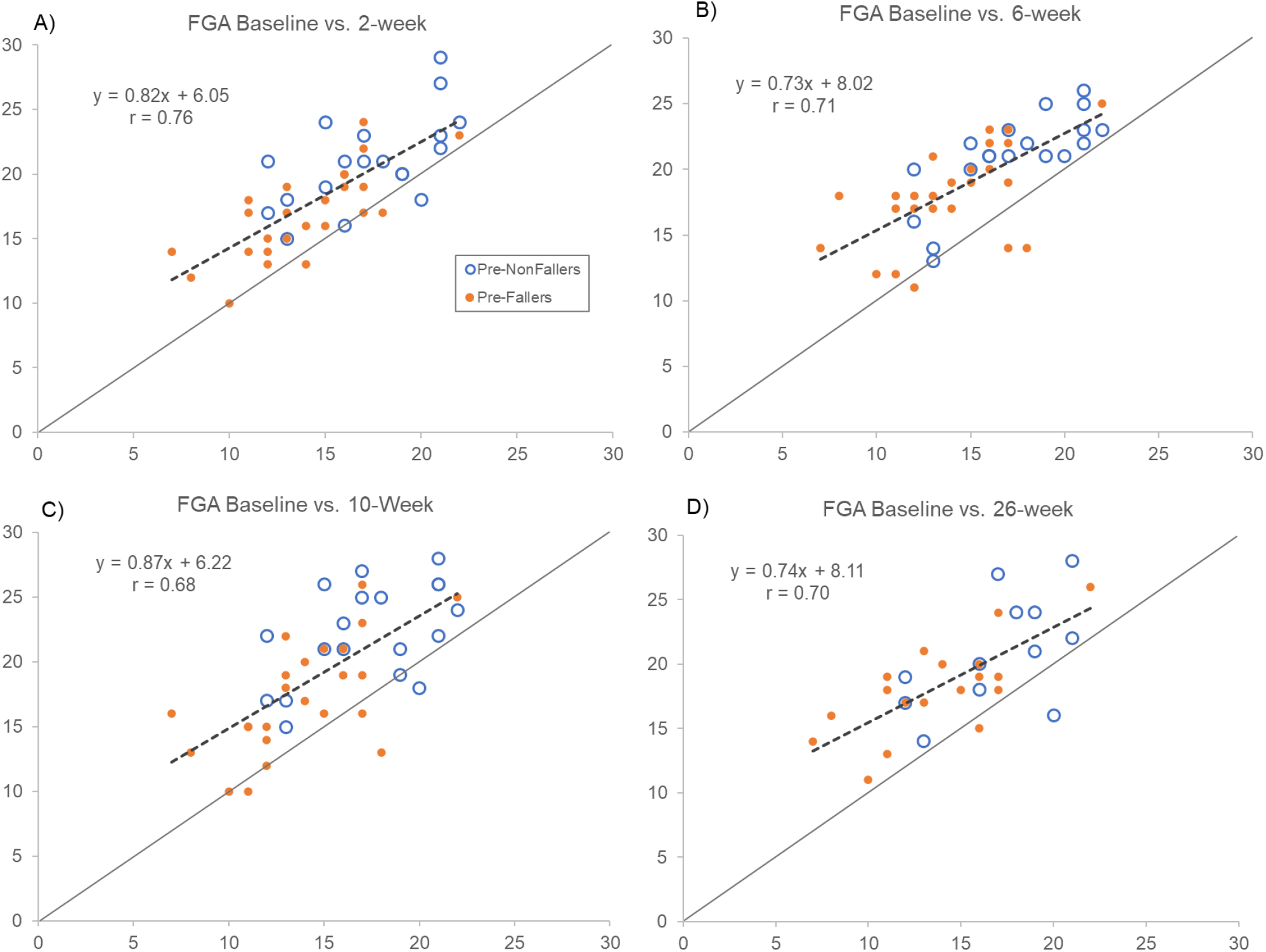
Comparing FGA scores at baseline (horizontal axes) with assessments after 2, 6, 10, and 26 weeks (vertical axes) of device use. Open symbols represent pre-study non-fallers and filled ones are pre-study fallers. Dashed lines show regression line for the whole group. Forty-four participants completed in clinic outcomes testing up to 10 weeks (primary endpoint), and 30 participants completed the 26-week assessment in person. Scores above line of equality indicate improvements and below a decrement in FGA score compared to baseline scores. Notice that regression line slopes are less than 1 indicating slightly larger improvements in FGA score for those with lower baseline scores. Overall, improvements observed after 2 weeks of use appeared sustained throughout the 26 weeks of use.

**Table 3.** Clinical outcomes and Rydel-Seiffer vibration sensation screening for the 30 individuals who were able to complete all in-person assessments at baseline and after 26 weeks as well as the subgroups of Pre-Fallers and Pre-NonFallers. Statistical significance is indicated in **bold**. Bonferroni’s adjustment of significance levels for correlated measures was applied (Uitenbroek 1997). Values in (italics) indicate Pearson’s correlation coefficient followed by the adjusted significance level required for an overall significance of 0.05 as marked with *. Cohen’s drm indicates effect size for change between baseline and the primary endpoint 10 weeks where 0.2 is represents a small effect, 0.5 a medium effect, and 0.8 a large effect. Values in parenthesis show 95% confidence interval. “estimate the effect size for single-group pretest-posttest designs” (Morris and DeShon 2002)

| All | Baseline Mean (SD)<br>n=30 | 26-week Mean (SD)<br>n=30 | p-level | Cohen's $d_{rm}$<br>(95% CI's) |
| --- | --- | --- | --- | --- |
| FGA Score | 15.0 (3.9) | **19.2 (4.1)**<br>(0.69) 0.036* | **<0.00001** | **1.38 (0.82-1.94)** |
| Gait Speed Self-selected (m/s) | 0.89 (0.26) | **0.97 (0.28)**<br>(0.80) 0.040* | **0.02** | **0.49 (-0.03-1.0)** |
| Gait Speed Fast (m/s) | 1.30 (0.41) | 1.37 (0.42) | 0.07 | n/a |
| TUG (s) | 13.2 (4.6) | 12.3 (3.4) | 0.20 | n/a |
| 4-Stage Balance Test (s) | 25.6 (7.2) | **28.4 (5.7)**<br>(0.65) 0.034* | **<0.01** | **0.47 (-0.05-0.98)** |
| R/L 1st MTP Joint | 2.8 (2.5) | 2.2 (2.4) | 0.07 | n/a |
| R/L Lateral Malleolus | 3.8 (2.2) | **3.2 (2.1)**<br>(0.60) 0.032* | **0.032** | **0.32 (-0.19-0.83)** |
| R/L Patella | 4.0 (2.2) | 4.0 (1.8) | 0.91 | n/a |
| Pre-Fallers | Baseline Mean (SD)<br>n=18 | 26-week Mean (SD)<br>n=18 | p-level | Cohen's $d_{rm}$<br>(95% CI's) |
| FGA Score | 13.7 (3.7) | **18.1 (3.6)**<br>(0.72) 0.037* | **<0.0001** | **1.58 (0.83-2.33)** |
| Gait Speed Self-selected (m/s) | 0.82 (0.25) | 0.90 (0.29) | 0.12 | n/a |
| Gait Speed Fast (m/s) | 1.13 (0.33) | 1.23 (0.37) | 0.07 | n/a |
| TUG (s) | 14.5 (5.0) | 13.0 (3.9) | 0.14 | n/a |
| 4-Stage Balance Test (s) | 23.5 (6.5) | **27.7 (5.4)**<br>(0.69) 0.037* | **<0.02** | **0.82 (0.14-1.50)** |
| R/L 1st MTP Joint | 2.9 (2.4) | **1.8 (2.0)**<br>(0.55) 0.03* | **0.005** | **0.58 (-0.09-1.24)** |
| R/L Lateral Malleolus | 3.75 (2.1) | 3.25 (1.9) | 0.07 | n/a |
| R/L Patella | 4.2 (2.0) | 4.0 (1.9) | 0.64 | n/a |
| Pre-NonFallers | Baseline Mean (SD)<br>n=12 | 26-week Mean (SD)<br>n=12 | p-level | Cohen's $d_{rm}$<br>(95% CI's) |
| FGA Score | 17.0 (3.3) | **20.8 (4.3)**<br>(0.56) 0.031* | **0.004** | **1.23 (0.36-2.10)** |
| Gait Speed Self-selected (m/s) | 1.00(0.25) | 1.08 (0.25) | 0.06 | n/a |
| Gait Speed Fast (m/s) | 1.56 (0.40) | 1.59 (0.42) | 0.63 | n/a |
| TUG (s) | 11.1 (3.1) | 11.3 (2.5) | 0.78 | n/a |
| 4-Stage Balance Test (s) | 28.7 (7.4) | 29.5 (6.1) | 0.65 | n/a |
| R/L 1st MTP Joint | 2.5 (2.8) | 2.7 (2.8) | 0.76 | n/a |
| R/L Lateral Malleolus | 3.7 (2.5) | 3.1 (2.4) | 0.21 | n/a |
| R/L Patella | 3.7 (2.5) | 4.0 (1.7) | 0.68 | n/a |

The Pre-F cohort (n=18) improved their mean FGA score from 13.7 at baseline to 18.1 at 26 weeks (p<0.0001, Table 3) representing a large effect size (Cohen’s drm=1.58). Their observed increases in self-selected gait speed from 0.82 m/s to 0.90 m/s and from 1.13 m/s to 1.23 m/s for fast gait speed did not reach statistical significance (p=0.12 and p=0.07, respectively). The Pre-F group showed a decrease in vibration sensation at the first MTP joint (mean 2.9 to 1.8, p<0.005) and at the lateral malleolus that did not reach statistical significance (mean 3.75 to 3.25, p=0.07).

The Pre-NF group (n=12) increased their mean FGA score from 17.0 at Baseline to 20.8 at 26 weeks (p<0.004, Cohen’s drm=1.23, Table 3). A small increase in self-selected gait speed from 1.00 m/s to 1.08 m/s did not reach statistical significance (p=0.06). Other clinical outcomes, including Rydel-Seiffer tuning fork sensitivity testing scores, remained unchanged at 26 weeks compared to baseline (Table 3).

The mean values of all in-person outcomes across the 26 weeks of device use for all participants are depicted in Figure 4 and are shown separately for the Pre-F and the Pre-NF groups. In general, the behavior over time of the two subgroups was qualitatively similar, although the Pre-NF group consistently performed better than the Pre-F cohort.

**Figure 4.**
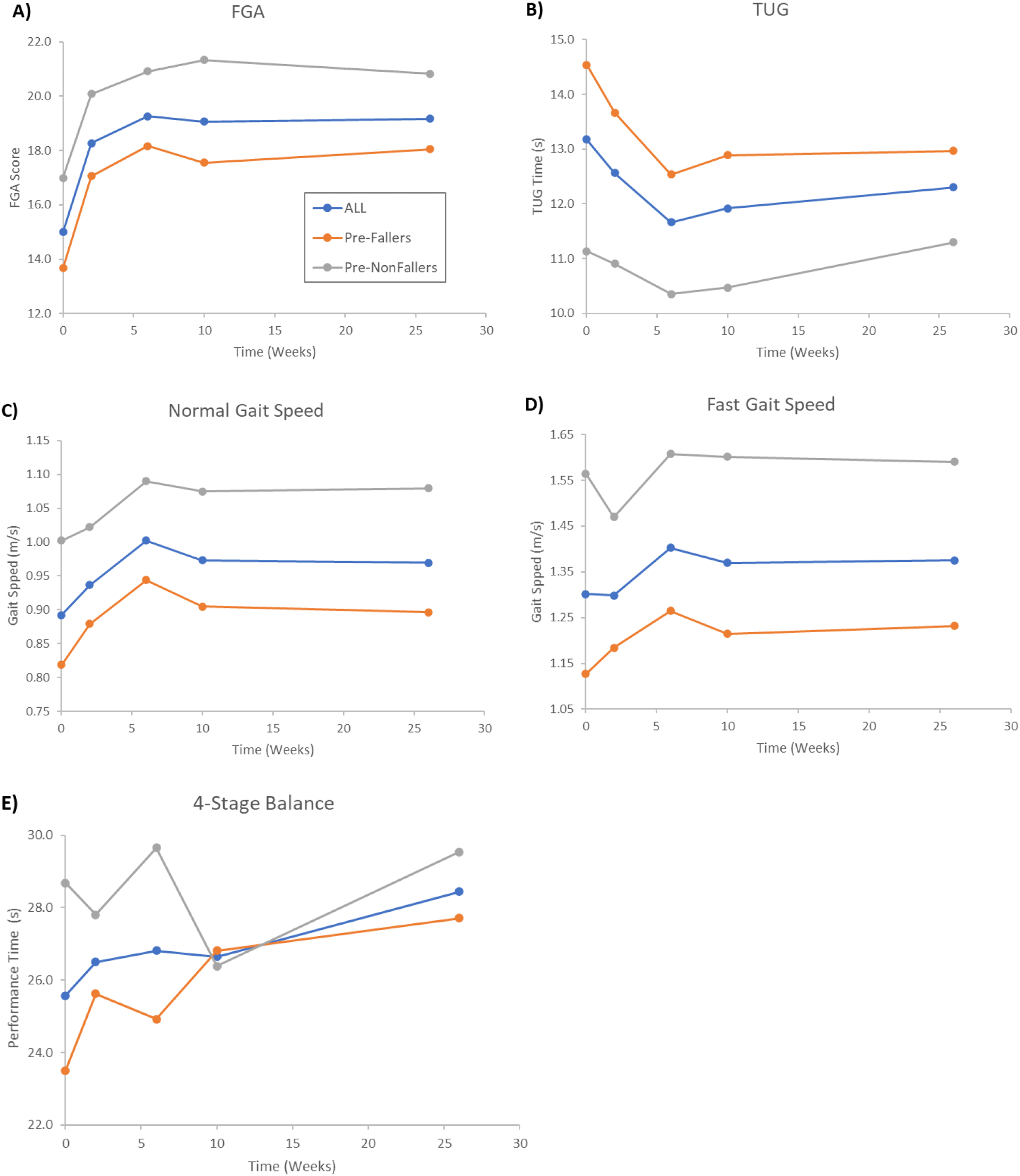
Averages of clinical outcomes across the in-person assessments at baseline (0), 2, 6, 10, and 26 weeks for the 30 participants (blue lines and symbols) who were able to be tested in clinic after 26 weeks of device use. Grey symbols and lines represent the pre-study non-fallers (n=12 of 30,) and orange lines and symbols represent pre-study fallers (n=18 of 30). Notice that most improvements appeared to peak after 6-10 weeks of device use, followed by a leveling off until 26 weeks.

### 3.4 Self-reported Outcomes, Device Use and ABC to FGA ratios

Self-reported outcomes for the 44 participants who completed assessments through 26 weeks are shown in Table 4. Across all participants, a slight but statistically significant increase was seen in PROMIS Ability to Participate scores after 26 weeks of device use (49.9 to 52.6, p<0.05). All baseline values were maintained at 26 weeks for the Pre-F group while we noted an increase in the PROMIS Satisfaction with Social Roles scores (49.9 to 55.5, p<0.006) for the Pre-NF group.

**Table 4.** Results from self-reported outcomes for the 44 individuals who completed all assessments. Data are shown separately for the group as a whole and for Pre-Fallers (having reported fallen in the prior 6 months) and Pre-NonFallers (no falls reported in the prior 6 months).

|  | **Baseline Mean (SD)**<br>**n=44** | **26-week Mean (SD)**<br>**n=44** | **p-level** |
| --- | --- | --- | --- |
| ABC-Score (%) | 62.9 (16.2) | 65.1 (14.8) | 0.39 |
| VADL Mean Score | 3.70 (1.06) | 3.46 (1.10) | 0.08 |
| VAS Score (0-10) | 2.9 (2.1) | 2.8 (2.2) | 0.66 |
| PHQ-9 | 4.7 (4.0) | 3.9 (4.2) | 0.14 |
| Pain Interference PROMIS® 6b | 50.9 (8.1) | 51.1 (8.3) | 0.88 |
| Satisfaction Social Roles PROMIS® 8a | 49.8 (7.0) | 51.9 (8.3) | 0.075 |
| Ability to Participate PROMIS® 8a | 49.9 (7.4) | 52.6 (8.6) | **<0.05** |
| **Pre-Fallers (n=25)** | **Baseline Mean (SD)** | **26-week Mean (SD)** |  |
| ABC-Score (%) | 57.8 (13.9) | 59.1 (14.4) | 0.73 |
| VADL Mean Score | 4.14 (1.00) | 3.95 (0.79) | 0.32 |
| VAS Score (0-10) | 3.0 (2.1) | 3.1 (1.9) | 0.76 |
| PHQ-9 | 5.7 (4.7) | 5.1 (4.5) | 0.22 |
| Pain Interference PROMIS® 6b | 52.7 (8.0) | 52.9 (7.3) | 0.86 |
| Satisfaction Social Roles PROMIS® 8a | 49.8 (6.7) | 49.2 (6.8) | 0.69 |
| Ability to Participate PROMIS® 8a | 48.4 (7.1) | 50.0 (7.1) | 0.12 |
| **Pre-NonFallers (n=19)** | **Baseline Mean (SD)** | **26-week Mean (SD)** |  |
| ABC-Score (%) | 69.6 (17.1) | 73.0 (12.0) | 0.34 |
| VADL Mean Score | 3.11 (0.86) | 2.81 (1.00) | 0.14 |
| VAS Score (0-10) | 2.8 (2.2) | 2.4 (2.4) | 0.35 |
| PHQ-9 | 3.3 (2.9) | 2.4 (3.5) | 0.35 |
| Pain Interference PROMIS® 6b | 48.5 (8.1) | 48.6 (9.2) | 0.96 |
| Satisfaction Social Roles PROMIS® 8a | 49.9 (7.7) | 55.5 (8.9) | **0.006** |
| Ability to Participate PROMIS® 8a | 51.8 (7.5) | 55.0 (10.0) | 0.18 |

Participants documented their weekly device use in a calendar and were asked to report it back during phone calls at weeks 14, 18, 22 and 26. Their average reported weekly device use was 5.1±0.4 days. Participants reporting that they used the device weekly either “Every Day” or “At least 5 Days” was 71.8±10.5%. An average of 94.8±3.3% of reporting participants stated they used the device “1-2 Days” or more per week.

We also sought to monitor participants’ self-perceived balance confidence (Lajoie and Gallagher 2004) in relation to their assessed gait function performance over time in the trial by measuring changes in the ABC to FGA ratio, which is shown in Figure 5. Figure 5 illustrates two observations. First, the ABC/FGA ratio for the overly confident individuals gradually decreased from high values at baseline to week 6 when the ratio essentially aligned with the low self-confidence participants, who maintained a consistent ABC/FGA ratio of about 3.5 throughout the 26 week period of the trial (Figure 5, left y-axis). Second, both groups of participants increased their FGA scores in a similar fashion although the overly confident participants showed higher levels of improvement in their FGA scores (Figure 5, right y-axis).

**Figure 5.**
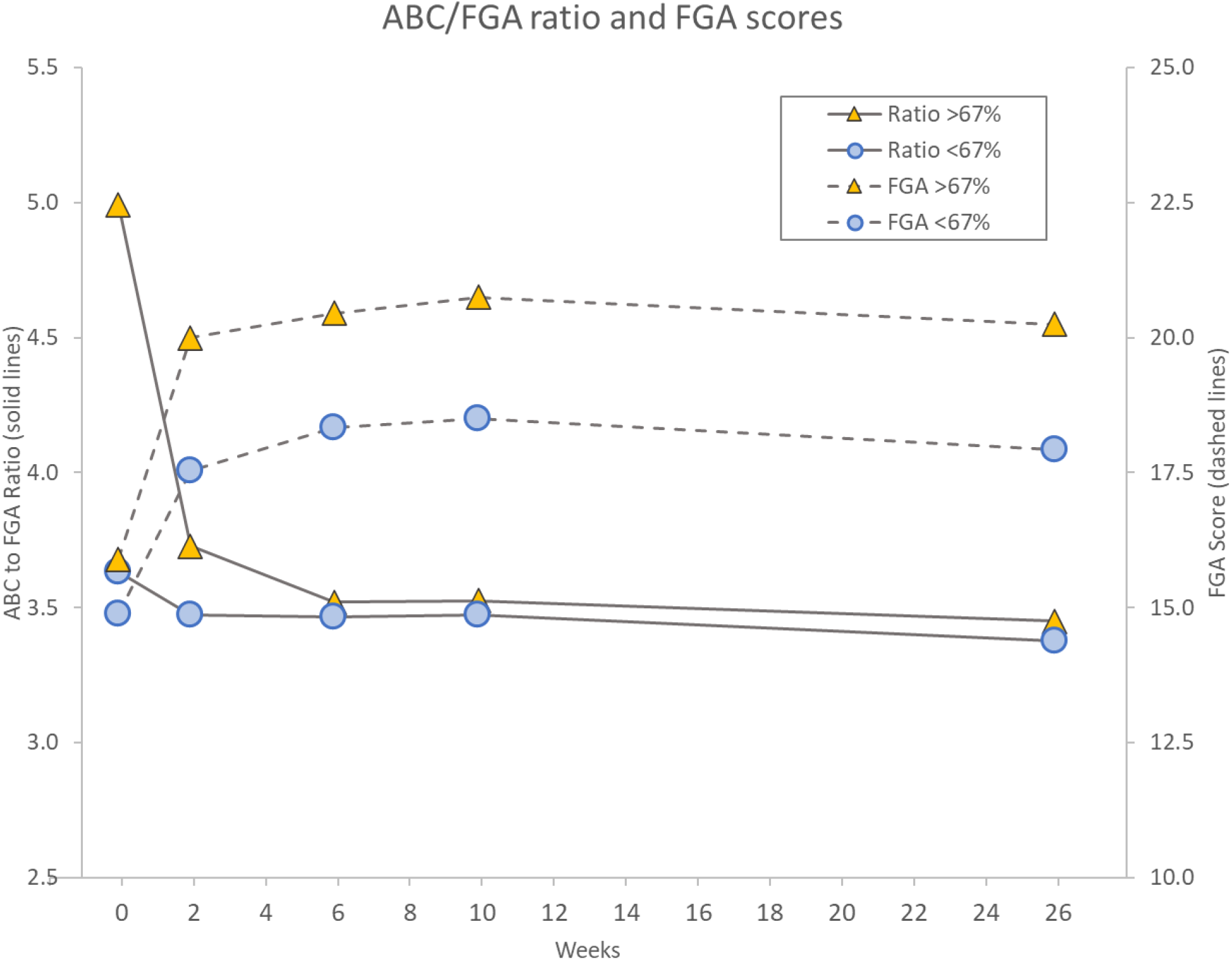
Illustration of changes in ABC/FGA ratio (left axis, solid lines), an indication of self-perceived balance confidence in relation to externally observed gait function performance, for participants with baseline ABC scores >67% (high balance-confidence, orange triangles) and those with ABC scores ≤67% (low balance-confidence, blue circles). The associated changes over time in FGA scores for the same two groups are also shown (right axis, dashed lines). Notice that both groups had similar FGA scores at baseline, which led to a dramatic difference in the ABC/FGA ratio at baseline. Over time, the ratio for the high balance-confidence participants gradually decreased and aligned with the low balance-confidence group at ∼3.5, mainly due to an increased FGA score and a maintained ABC score. The low balance-confidence group maintained an ABC/FGA ratio ∼3.5 throughout the 26 weeks associated with a proportional increase in ABC and FGA scores. Interestingly, the high balance-confidence group showed a higher increase in FGA score than the low balance-confidence group.

### 3.5 Falls Data

Falls were reported throughout the 26-week period for all 44 participants and separately for the Pre-F (n=25) and Pre-NF (n=19) groups, respectively (Table 5). The 44 participants reported a total of 53 falls over 6 months prior to participating in the trial while 39 falls were documented during the 26 weeks of the trial, corresponding to a pre-study mean fall rate of 6.7 falls/1000 patient days (median=5.6 falls/1000 patient days) and a post study mean fall rate of 4.8 falls/1000 patient days (median=0 falls/1000 patient days). Wilcoxon Signed Rank test was used to compare pre- and post-study fall rates since the data was not normally distributed (Shapiro-Wilk W-statistic=0.82, p<0.0001). Across all participants, the median of the post-study fall rate was lower than the pre-study fall rate (Table 5, p=0.044), reflecting a 28% decrease in fall rate. Of the 44 participants, 25 had fallen in the past six months (Pre-F); and after 26 weeks, 20 of the 44 participants had reported falling. Overall, 31 of the 39 falls required no treatment while eight falls (∼20%) required treatment with four of those (∼10%) causing severe injury (two fractures) (Table 5).

**Table 5.** Parameters related to falls and fall risk assessed at baseline and at 26 weeks. All 44 individuals were asked to report their falls throughout the 26-week period. Data for Pre-Fallers and Pre-NonFallers are presented separately. Fall rates are reported in number of falls per 1000 patient days. Fall rates were not normally distributed (Shapiro–Wilk W-stat =0.82). *Wilcoxon Signed Rank test.

| **All (n=44)** | **Baseline (SD)** | **26 Weeks (SD)** | **p-level** |
| --- | --- | --- | --- |
| #Falls (pre-6 mo & in study, n=44) | 53 | 39 | n/a |
| Fall Rate Mean (#/1000 patient days, pre-6mo & in study, n=44) | 6.7 (7.7) | 4.8 (7.7) | n/a |
| Fall Rate Median | 5.6 | 0 | **0.044\*** |
| #Fallers (pre-6mo & at 6mo in study, n=44) | 25 | 20 | 0.28 |
| Falls no treatment sought | n/a | 31 | n/a |
| Falls treatment sought + severe injury | n/a | 4+4 | n/a |
| **Pre-Fallers (n=25)** |  |  |  |
| #Falls (pre-6 mo & in study, n=25) | 53 | 31 | n/a |
| Fall Rate Mean (#/1000 patient days, pre-6mo & in study, n=25) | 11.8 (6.7) | 6.7 (9.5) | n/a |
| Fall Rate Median | 11.1 | 5.0 | **0.0043\*** |
| #Fallers (pre-6mo & at 6mo in study, n=25) | 25 | 13 | **<0.0001** |
| Falls no treatment sought | n/a | 25 | n/a |
| Falls treatment sought + severe injury | n/a | 3+3 | n/a |
| **Pre-NonFallers (n=19)** |  |  |  |
| #Falls (pre-6 mo & in study, n=19) | 0 | 8 | n/a |
| Fall Rate Mean (#/1000 patient days, pre-6mo & in study, n=19) | 0 (0) | 2.3 (3.3) | n/a |
| Fall Rate Median | 0 | 0 | **0.023\*** |
| #Fallers (pre-6mo & at 6mo in study, n=19) | 0 | 7 | **<0.0001** |
| Falls no treatment sought | n/a | 6 | n/a |
| Falls treatment sought + severe injury | n/a | 1+1 | n/a |

The 25 participants in the Pre-F cohort reported 31 falls after 26 weeks compared to 53 falls pre-study corresponding to a pre-study fall rate of 11.8 falls/1000 patient days (median=11.1), which decreased to 6.7 falls/1000 patient days at 26 weeks (median=5.0), a 43% decrease in fall rate (p=0.0043). Of the 25 Pre-F participants, 12 did not fall during the study, a 48% statistically significant decrease in number of fallers (p<0.0001). Of the 31 falls experienced by the Pre-F cohort, six (∼20%) required treatment (Table 5). Three of the four falls that led to severe injury occurred in the Pre-F group.

Seven of the 19 Pre-NF participants reported falling during the trial (p<0.0001) leading to an increase in fall rate from zero to 2.3 falls/1000 patient days at 26 weeks that was statistically significant (p=0.023). A total of eight falls were reported by the seven Pre-NF participants who fell during the study (Table 5). Two of the eight falls in the group required treatment, and one fall led to a severe injury.

Figure 6 illustrates the cumulative sum of falls based on six-month pre-study fall reports from the participants and the falls recorded by the Pre-F cohort during the trial. The 53 pre-study falls were randomly distributed across the six-month pre-study period for illustration purposes since the exact time of their occurrence prior to the trial were not known (see Figure 6). Note that 53 falls over a period of six months corresponds to 53/180∼0.29 falls/day, which becomes the slope of the blue regression line in Figure 6, irrespective of the random distribution of the pre-study falls. The in-study falls are presented on the day they occurred according to participant reports (see Figure 6). The in-study decrease of falls quantified in Table 5 can be visually observed over time in Figure 6 as a lower slope for the best fit regression line (blue-dotted line pre-study vs. orange-dotted line in-study). It should be noted that the lower slope appears to begin after about 20 days of device use.

**Figure 6.**
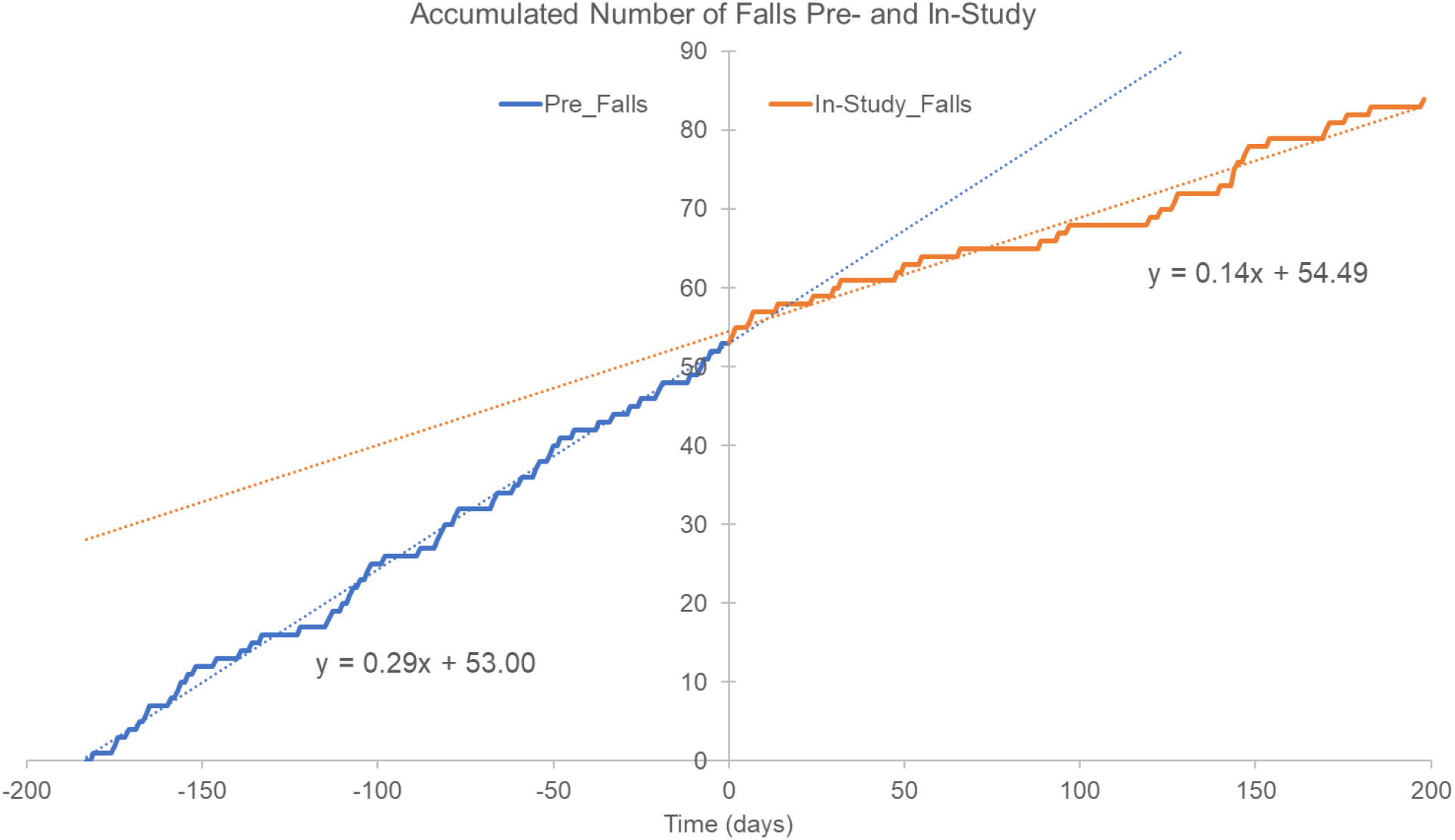
Accumulated number of falls reported six months pre-study (blue trace and blue dotted linear regression line) and falls documented in-study (orange trace and orange dotted regression line). The 53 falls reported for the prior six months would correspond to 0.29 falls/day (53/180∼0.29), which represents the slope of the regression line (blue trace). For illustration purposes, the 53 pre-study falls were randomly distributed across the six months since their exact time occurrence was unknown. In-study falls are shown as they occurred and were reported by participants throughout the 26 weeks. Notice how the rate of in-study falls appear to begin deviating from pre-study fall rate (slope of dotted blue line) after approximately 20 days of device use.

## 4 Discussion

### 4.1 Key Findings

Walkasins is a device that provides mechanical tactile stimuli related to foot pressure for individuals with PN and gait and balance problems. Overall, our findings from analyzing outcomes after 26 weeks of wearing Walkasins show that improvements in outcomes seen after 10 weeks of use are sustained longer term (Oddsson, Bisson et al. 2020). The FGA score was improved from 15.0 at baseline to 19.2 after 26 weeks of device use across all participants, a large effect size (Cohen’s drm=1.38) and an increase that is beyond the MCID for community-dwelling older adults (Beninato, Fernandes et al. 2014). Interestingly, these improvements were seen for individuals across the full range of baseline FGA scores, from the lowest of 7 to the highest of 22 (Figure 3) with a tendency to be higher for individuals in the lower FGA range. Furthermore, similar improvements were seen for the Pre-F and Pre-NF cohorts. Moreover, the 46% decrease in fall rate, compared to pre-study falls we reported after 10 weeks of use, was sustained at 43% after 26 weeks, an important observation further discussed below. Weekly device use continued to be high and similar to after 10 weeks (5.1 vs. 5.3 days/week, respectively (Oddsson, Bisson et al. 2020)).

All clinical outcomes improved compared to baseline, but changes in TUG and 4-Stage Balance did not reach statistical significance, likely because the study was underpowered for these measures (Table 3). Previous work has found that older individuals with diabetic PN walk slower than their healthy cohort (Menz, Lord et al. 2004), and older adults with peripheral sensory impairment show a 0.046m/s/yr. decline in gait speed (Lipsitz, Manor et al. 2018). This decrease is more than 3.5 times higher than declines reported in healthy aging (Buracchio, Dodge et al. 2010). With this in mind, it is encouraging to report a statistically significant 0.08m/s increase in gait speed gait speed across all participants in this trial, which is beyond a small meaningful change (0.05m/s) and close to the range for a substantial meaningful change (0.10m/s) (Perera, Mody et al. 2006).

Participants had a decrease in sensitivity to vibration, an overall small decrease at the lateral malleolus and a larger change at the MTP joint in the Pre-Faller group. Such changes did not occur at the 10-week assessment (Oddsson, Bisson et al. 2020). This decrease in tuning fork vibration perception may indicate a progression of some participants’ PN and increased sensory loss. In spite of such changes, none of the participants stated that they were unable to perceive the stimuli provided by the device, suggesting they were still benefiting from its use as confirmed by improved clinical outcomes.

### 4.2 Balance Exercise Interventions and their Limitations

We can compare our findings to interventions using different forms of physical exercise in patients with PN since we see similar improvements in outcomes from simply wearing the device in the current trial. Several review studies conclude that balance training is “the most effective exercise intervention” (Ites, Anderson et al. 2011, Tofthagen, Visovsky et al. 2012, Streckmann, Zopf et al. 2014, Morrison, Simmons et al. 2018), while strength training in patients with PN appears to have less impact on balance (Streckmann, Zopf et al. 2014), likely due to lack of specificity related to balance (Oddsson, Boissy et al. 2007). Although such interventions may improve balance outcomes, effects are essentially compensatory since they do not address the root cause of the balance problem in these patients, namely their impaire plantar sensation due to PN. Furthermore, any exercise activity would need to be maintained on a regular basis long-term or benefits would gradually be lost (Melzer and Oddsson 2013, van Waart, Stuiver et al. 2015), sometimes referred to as the “use it or lose it” principle (Hart 2021). This phenomenon is problematic because individuals sometimes become sedentary due to health problems unrelated to their balance disorders. Likewise, long-term compliance with exercise interventions is a challenge, due to motivation or lack of safety resources, and has been reported to be low with over half of participants not being compliant (Sluijs, Kok et al. 1993, Alexandre, Nordin et al. 2002), likely causing diminished benefits of the exercise intervention.

The improvements in clinical outcomes we report here, were gained from a single, brief training visit on the use of the sensory prosthesis and then simply by wearing it during regular daily activities. By comparison, obtaining the benefits of an exercise intervention requires the presence and engagement by the individual, at multiple therapy visits. In fact, during the first 10 weeks of use, we prohibited participants from joining in any physical therapy or balance-related exercise activities to help isolate the effect of the device (Oddsson, Bisson et al. 2020). Knowing, however, that such interventions may help improve gait and balance function (Tofthagen, Visovsky et al. 2012), we felt it would potentially be unsafe and even unethical to prevent participation in those activities beyond the primary endpoint at 10 weeks. Therefore, to control for any potential effects of balance exercise interventions, we asked participants during follow-up phone calls at 14, 18, and 22 weeks whether they had begun any physical therapy for their balance problems. Not surprisingly, only one of the participants answered “yes” to this question, emphasizing the above-mentioned challenge with exercise engagement and compliance, and further suggesting that long-term use of the device was the main cause of improvement in clinical outcomes.

### 4.3 Exercise Dosing versus Daily Exposure from Sensory Prosthesis Use

These improvements in clinical outcomes are not surprising, because the mechanisms for improved gait and balance function from exercise interventions are different than the mechanisms for using a sensory prosthesis to replace an important component of somatosensory balance input, in this case from plantar mechanoreceptors (Meyer, Oddsson et al. 2004, Fallon, Bent et al. 2005, Strzalkowski, Peters et al. 2018). Traditional balance training activities stimulate existing sensorimotor integration by challenging function with a series of successively more difficult motor tasks. This kind of treatment gradually improves performance based on important and well-known principles of training (Oddsson, Boissy et al. 2007). Such improvement may be attributable to training of specific splinter skills, and some improvement may be due to compensation for balance issues from impaired plantar sensation or address other balance related impairments. Users of a sensory prosthesis, however, receive new functional sensory stimuli on a nearly continuous basis that are highly specific to standing and walking and replace their impaired sense; and these new stimuli provide sensory information that the central nervous system expects to receive from the lower extremities during such activities (Guertin 2012, Clark 2015).

To analyze this contrast further, a research trial of traditional balance training interventions typically provides one-hour balance stimulation sessions about 2-3 times per week for 3-6 months e.g. (Shumway-Cook, Gruber et al. 1997, Wolf, Feys et al. 2001, Wolf, Sattin et al. 2003, Li, Harmer et al. 2005, Li and Manor 2010, Melzer and Oddsson 2013, Manor, Lough et al. 2014). Assuming 100% compliance, this example would add up to an overall exercise dose of no more than 72 hours of focused therapeutic balance stimulation over six months. In a typical therapy practice, the dose would more likely be in the range of 8-40 hours, for several reasons (HS Cohen, EdD, OTR, personal communication). Patients sometimes miss visits due to schedule conflicts. Patients may shorten the total length of treatment either because they feel they have reached their maximum level of improvement, because they dislike the hassle of taking the time for the therapy visit, or because they cannot get transportation. In addition, some insurance programs may refuse to pay for more than a limited number of therapy visits. In the case of exposure to balance stimuli from a sensory prosthesis, however, participants used the device on average 36.1 hours/week with more than 80% using the device over 21 hours/week (Oddsson, Bisson et al. 2020). Over 26 weeks this use would amount to a range from around 500 to almost 1000 hours of specific gait and balance stimulation, essentially an order of magnitude higher than a regular balance training intervention. Furthermore, the device is intended to be used on a continuous basis, which furthers compliance.

Participants may not be physically active and receiving sensory stimuli the entire abovementioned time. However, they will regularly experience hundreds of specific balance stimuli/hour that are intimately integrated into their regular standing and walking activities and that are likely to cause related changes in balance behavior. Over time, this feedback may lead to a reweighting of sensory information that is relevant for balance function (Sienko, Seidler et al. 2018), as has previously been suggested in older adults performing in-home balance training using vibrotactile sensory stimulation related to trunk tilt (Noohi, Kinnaird et al. 2017, Bao, Carender et al. 2018) further supported by recently reported changes in brain connectivity (Bao, Noohi et al. 2022).

Interestingly, early observations from a pilot study of five participants in the cohort studied here, who completed 26 weeks of device use, show neuroplastic changes in brain network connectivity related to postural control and balance that were associated with improved changes in FGA scores (Hsu, Iloputaife et al. 2021). This finding indicates a direct effect of the sensory prosthesis initiating plastic changes related to sensorimotor interaction and postural control. Related observations on sensorimotor neuroplastic effects associated with peripheral afferent activity have previously been made following a novel amputation strategy, agonist-antagonist myoneural interface, intended to maintain neuromuscular communication of the lost limb (Srinivasan, Tuckute et al. 2020). This surgical technique intends to promote proprioceptive feedback and cause central sensorimotor plastic changes that may facilitate control of a prosthetic limb (Srinivasan, Tuckute et al. 2020). In a similar fashion, the current non-invasive sensory prosthesis is externally providing new tactile afference, essentially through the same dermatomes that signaled plantar pressure information from mechanoreceptors that have become functionally deafferented due to PN (Fallon, Bent et al. 2005, Koehler-McNicholas, Danzl et al. 2019, Vaughan 2021).

### 4.4 ABC to FGA Ratios, Self-Reported Outcomes and Device Use

On the ABC, 67% is the cut-off for high-fall risk (Lajoie and Gallagher 2004). We previously reported that participants with baseline ABC scores below 67% improved their ABC scores after 10 weeks of device use, whereas those with ABC scores above 67% did not, although both subgroups improved their FGA scores (Oddsson, Bisson et al. 2020). Since these two categories of participants had similar baseline FGA scores, the discrepancy in ABC scores led to seemingly high baseline ABC to FGA ratios in participants with high ABC scores (Oddsson, Bisson et al. 2020) (Figure 5). We posited that the high ratio individuals were either too confident or simply unaware of their abilities or impairments, a construct that aligns with a proposed multifactorial causation model for falls emphasizing the importance of a “Realistic Appraisal of One’s Own Abilities”, (Hadjistavropoulos, Delbaere et al. 2011). This ratio indicates the degree of internal self-perception of balance capability (ABC score) in relation to the externally observed performance (FGA score). A ratio of around 3.3 would be “expected” based on the maximum scores of the two scales (Powell and Myers 1995, Wrisley and Kumar 2010). We further noted that after six weeks of device use, the overly confident participants appeared to “normalize” their self-perception indicating a more sensible perception of their abilities and align with those holding more expected ABC scores. This change was mainly due to an improvement in their FGA score and a maintained or slightly decreased ABC score.

We show here that this observation is preserved after 26 weeks of device use (Figure 5). This change in self confidence is important because it is a fall risk factor. As the data suggest, this ratio is a modifiable risk factor. Therefore, the ratio can and should be measured during studies of balance interventions.

Furthermore, the current data (Figure 5) shows that the high-confidence individuals appear to improve their FGA scores more than the low confidence individuals. Since the baseline FGA scores were similar for these two cohorts, it could be argued that the higher balance-confidence individuals may have “naturally” challenged themselves more in their daily activities, triggering a larger increase in FGA score. Correspondingly, the low balance-confidence individuals may have “explored” their improved functional capabilities less than the high confidence group leading to a less beneficial long-term effect. We postulate that individuals with low balance confidence would potentially benefit further from a guided interaction through physical and occupational therapy or other means, whereas the high confidence individuals may be better at “self-coaching” their renewed capabilities. Consequently, to provide an individually optimized rehabilitation strategy for falls prevention and self-efficacy, clinicians should monitor outcomes that measure both balance outcomes and balance confidence, similar to the ratio we have used to allow this construct to be investigated further (Hadjistavropoulos, Delbaere et al. 2011, Soh, Tan et al. 2021).

Additional observations related to self-report measures were in line with our previous reporting (Oddsson, Bisson et al. 2020), overall showing marginal changes and likely being less clinically meaningful since they were near the average for the US population (Hahn, DeWalt et al. 2014, Askew, Cook et al. 2016, Hahn, Beaumont et al. 2016, Hahn, Kallen et al. 2016). The continued high level of device use, on average more than five days a week and over 70% of participants using it every day or at least 5 days/week, suggests that users find the device helpful.

### 4.5 Falls Data

One of the most important findings from this trial is the 43% decrease in fall rate for participants with a fall history following 26 weeks of device use. We reported an encouraging 46% decrease already after 10 weeks of use (Oddsson, Bisson et al. 2020), while appreciating that 10 weeks would not be long enough for a relevant comparison to six months of pre-study data. Consequently, the similar statistically significant decrease after 26 weeks provides a stronger argument for a meaningful decrease in falls. In fact, even including the pre-study non-fallers in the analysis showed a statistically significant decrease in fall rate over 26 weeks of device use (Table 5). Note, however, that fall rate was not a primary outcome in this trial. In fact, we gathered pre-study falls data primarily as a means of describing our study population, and we monitored falls during the trial as a subset of adverse events. Thus, we enrolled participants at high fall risk based on their FGA scores, irrespective of whether or not they had fallen in the past. This procedure allowed us to conduct a post-hoc analysis and investigate any trend in the data indicating an actual decrease in falls. A comparison with historical self-report data on prior falls is far from ideal as compared to a parallel arm trial with a control group, but other investigators have found that asking individuals to recall past falls commonly leads to an underreporting of actual falls; even recalling the past 3 months led to 25% underreporting of falls (Hannan, Gagnon et al. 2010), and 26% did not recall falls over the prior six months (Cummings, Nevitt et al. 1988). Under such circumstances, the 43% decrease in falls reported here probably underestimates the real effect.

The data on injuries reported from falls are probably clinically significant. Previous studies have reported that 65% of older individuals with PN fell over a year with 30% reporting an injury from a fall (DeMott, Richardson et al. 2007). In a study of individuals with chemotherapy-induced PN, over 40% experienced falls, and more than 40% of falls resulted in an injury, two-thirds of which were fractures (Komatsu, Yagasaki et al. 2019). In this study, 57% of participants reported falling in the six months prior to the study while 45% fell during six months of the trial. Eight of the 39 falls during the trial (20.5%) led to injuries where treatment was sought, half of those considered serious. Consequently, both fall rate and injury rate are lower than previous reports of similar populations (DeMott, Richardson et al. 2007, Komatsu, Yagasaki et al. 2019).

### 4.6 Study Limitations

This study had several limitations, as we have previously outlined (Oddsson, Bisson et al. 2020). It was an unblinded, single arm trial. Our decision to conduct a single arm trial was based on several factors, including previous results from an in-clinic randomized control cross-over trial showing improved FGA scores when using the device turned on compared to off (Koehler-McNicholas, Danzl et al. 2019). In addition, data from the first-in-human, long-term use study showed remarkable improvements in clinical outcomes in a patient with PN using the device for a year following over five months of balance physical therapy with limited improvement (Wrisley, McLean et al. 2021).

This individual continues to use the device daily, now for more than four years. Also, blinding participants in a study using this kind of intervention is challenging and may not really be viable unless some form of deception is used since a requirement to use the device is perceiving the tactile stimuli (Oddsson, Bisson et al. 2020). Consequently, the most feasible placebo treatment for a control group would likely be wearing a device that is non-functional.

An important strength of the trial includes our real-life inclusion/exclusion criteria that were essentially the same as the requirements for receiving a prescription for the device outside of a research trial. These requirements are aligned with the intentions of a pragmatic clinical trial to advance applicability of study findings (Patsopoulos 2011, Gill, Bhasin et al. 2021). Only 7% of participants (6 of 85) were excluded due to medical circumstances (Figure 2). We wanted the participants to realistically reflect patients who are seen in the clinic, and we did not try to screen for only those individuals who may be the best responders. In fact, our participants reported an average of 8.3 chronic conditions, had multiple fall-risk factors, and polypharmacy (mean of 8 subscription medications).

Furthermore, the long-term benefits we report here combined with the participants not receiving any useful feedback about their outcomes or systematic encouragement during their assessments and knowing that only one individual decided to participate in a balance therapy intervention, decreases the chance these effects are placebo (Finniss, Kaptchuk et al. 2010, Enck, Bingel et al. 2013, Coste and Montel 2017). A review of placebo effects in 47 randomized control trials found large effects in subjective outcomes and small effects in clinical outcomes, which is the opposite of what we report here, further suggesting effects are due to device use. Additional strengths include our use of recommended standardized objective clinical outcome measures (Moore, Potter et al. 2018) and the involvement of multiple clinical sites from different regions using different evaluators.

## 5 Conclusion

Patients with peripheral neuropathy who have gait and balance problems with a high risk of falls including a history of falls improve their walking balance and decrease their fall rates from long-term use of a wearable sensory prosthesis.

## Data Availability

Raw data supporting the conclusions of this article will be made available to qualified researchers by the authors, without undue reservation

## 6 Acknowledgments

We thank Lori Danzl, Alexandria Lloyd, Christine Olney (Minneapolis Department of Veterans Affairs Health Care System, Minneapolis, MN), Jackie Geiser, Tien Dat Nguyen, Delorianne Sander (M Health Fairview, Minneapolis, MN), Nathan Silver (Baylor College of Medicine, Houston, TX), Wanting Yu (Hinda and Arthur Marcus Institute for Aging Research, Hebrew SeniorLife, Roslindale, MA) for providing study coordination, participant recruitment and data gathering activities. We thank Amy Gravely, MA, for input on statistical analysis and Annette Xenopoulos-Oddsson (University of Minnesota, MN,), Dr. Lee Newcomer, and Dr. Steven Stern for reviewing drafts of the manuscript. The manuscript was published as a MedRxiv preprint.

## 7 Contribution to the Field Statement

Falls in older adults continues to be a large problem with few solutions. Falls are associated with enormous costs to society, individual suffering, and loss of quality of life. Peripheral neuropathy, leading to nerve damage, is recognized as an independent risk factor for falls and is often related to diabetes or chemotherapy treatment or is of unknown etiology. A common symptom involves numbness and sensory loss in the feet that can directly cause falls. Here we report data after extended long-term daily use of a wearable sensory prosthesis (Walkasins^®^, RxFunction Inc., MN, USA) from the multi-site clinical trial, walk2Wellness. The device provides directional tactile stimuli around the ankle during standing and walking activities, reflecting changes in foot pressure related to balance. Patients can then “feel” their feet in contact with the ground and notice body sway during standing because the device replaces related aspects of their impaired afferent nerve function. Data after 26 weeks of use shows that previously reported improvements in clinical outcomes after 10 weeks of use are sustained. In addition, we can now report a decrease in fall rate in individuals who reported falling six months prior to participating in the study.

## References

Alexandre, N. M., M. Nordin, R. Hiebert and M. Campello (2002). “Predictors of compliance with short-term treatment among patients with back pain.” Rev Panam Salud Publica 12(2): 86–94.

Algina, J, H. J. Keselman and R. D. Penfield (2005). “Effect Sizes and their Intervals: The Two-Level Repeated Measures Case.” Edu Psych Meas 65(April): 241–258.

Askew, R. L., K. F. Cook, D. A. Revicki, D. Cella and D. Amtmann (2016). “Evidence from diverse clinical populations supported clinical validity of PROMIS pain interference and pain behavior.” J Clin Epidemiol 73: 103–111.

Bao, T., W. J. Carender, C. Kinnaird, V. J. Barone, G. Peethambaran, S. L. Whitney, M. Kabeto, R. D. Seidler and K. H. Sienko (2018). “Effects of long-term balance training with vibrotactile sensory augmentation among community-dwelling healthy older adults: a randomized preliminary study.” J Neuroeng Rehabil 15(1): 5.

Bao, T., F. Noohi, C. Kinnaird, W. J. Carender, V. J. Barone, G. Peethambaran, S. L. Whitney, R. D. Seidler and K. H. Sienko (2022). “Retention Effects of Long-Term Balance Training with Vibrotactile Sensory Augmentation in Healthy Older Adults.” Sensors (Basel) 22(8).

Beninato, M., A. Fernandes and L. S. Plummer (2014). “Minimal clinically important difference of the functional gait assessment in older adults.” Phys Ther 94(11): 1594–1603.

Bergen, G., M. R. Stevens and E. R. Burns (2016). “Falls and Fall Injuries Among Adults Aged ≥65 Years - United States, 2014.” MMWR Morb Mortal Wkly Rep 65(37): 993–998.

Buracchio, T., H. H. Dodge, D. Howieson, D. Wasserman and J. Kaye (2010). “The trajectory of gait speed preceding mild cognitive impairment.” Arch Neurol 67(8): 980–986.

Cavanagh, P. R., J. A. Derr, J. S. Ulbrecht, R. E. Maser and T. J. Orchard (1992). “Problems with gait and posture in neuropathic patients with insulin-dependent diabetes mellitus.” Diabet Med 9(5): 469–474.

CDC. (2017). “STEADI -Older Adult Fall Prevention.” Retrieved May 5, 2018, 2018, from https://www.cdc.gov/steadi/materials.html.

CDC. (2021). “Older Adult Fall Prevention.” Retrieved November 1, 2021, 2021, from https://www.cdc.gov/falls/facts.html.

Cesari, M., S. B. Kritchevsky, B. W. Penninx, B. J. Nicklas, E. M. Simonsick, A. B. Newman, F. A. Tylavsky, J. S. Brach, S. Satterfield, D. C. Bauer, M. Visser, S. M. Rubin, T. B. Harris and M. Pahor (2005). “Prognostic value of usual gait speed in well-functioning older people--results from the Health, Aging and Body Composition Study.” J Am Geriatr Soc 53(10): 1675–1680.

Clark, D. J. (2015). “Automaticity of walking: functional significance, mechanisms, measurement and rehabilitation strategies.” Front Hum Neurosci 9: 246.

Clark, D. J., E. A. Christou, S. A. Ring, J. B. Williamson and L. Doty (2014). “Enhanced somatosensory feedback reduces prefrontal cortical activity during walking in older adults.” J Gerontol A Biol Sci Med Sci 69(11): 1422–1428.

Cohen, H. S., K. T. Kimball and A. S. Adams (2000). “Application of the vestibular disorders activities of daily living scale.” Laryngoscope 110(7): 1204–1209.

Cohen, J. (1988). Statistical Power Analysis for the Behavioral Sciences. New York, NY, Routledge Academic.

Cohen, M. A., J. Miller, X. Shi, J. Sandhu and L. A. Lipsitz (2015). “Prevention Program Lowered The Risk Of Falls And Decreased Claims For Long-Term Services Among Elder Participants.” Health Aff (Millwood) 34(6): 971–977.

Coste, J. and S. Montel (2017). “Placebo-related effects: a meta-narrative review of conceptualization, mechanisms and their relevance in rheumatology.” Rheumatology (Oxford) 56(3): 334–343.

Cummings, S. R., M. C. Nevitt and S. Kidd (1988). “Forgetting falls. The limited accuracy of recall of falls in the elderly.” J Am Geriatr Soc 36(7): 613–616.

DeMott, T. K., J. K. Richardson, S. B. Thies and J. A. Ashton-Miller (2007). “Falls and gait characteristics among older persons with peripheral neuropathy.” Am J Phys Med Rehabil 86(2): 125–132.

Dixon, C. J., T. Knight, E. Binns, B. Ihaka and D. O’Brien (2017). “Clinical measures of balance in people with type two diabetes: A systematic literature review.” Gait Posture 58: 325–332.

Dumurgier, J., A. Elbaz, P. Ducimetière, B. Tavernier, A. Alpérovitch and C. Tzourio (2009). “Slow walking speed and cardiovascular death in well functioning older adults: prospective cohort study.” BMJ 339: b4460.

Enck, P., U. Bingel, M. Schedlowski and W. Rief (2013). “The placebo response in medicine: minimize, maximize or personalize?” Nat Rev Drug Discov 12(3): 191–204.

Fallon, J. B., L. R. Bent, P. A. McNulty and V. G. Macefield (2005). “Evidence for strong synaptic coupling between single tactile afferents from the sole of the foot and motoneurons supplying leg muscles.” J Neurophysiol 94(6): 3795–3804.

Finniss, D. G., T. J. Kaptchuk, F. Miller and F. Benedetti (2010). “Biological, clinical, and ethical advances of placebo effects.” Lancet 375(9715): 686–695.

Ganz, D. A. and N. K. Latham (2020). “Prevention of Falls in Community-Dwelling Older Adults.” N Engl J Med 382(8): 734–743.

Gill, T. M., S. Bhasin, D. B. Reuben, N. K. Latham, K. Araujo, D. A. Ganz, C. Boult, A. W. Wu, J. Magaziner, N. Alexander, R. B. Wallace, M. E. Miller, T. G. Travison, S. L. Greenspan, J. H. Gurwitz, J. Rich, E. Volpi, S. C. Waring, T. M. Manini, L. C. Min, J. Teresi, P. C. Dykes, S. McMahon, J. M. McGloin, E. A. Skokos, P. Charpentier, S. Basaria, P. W. Duncan, T. W. Storer, P. Gazarian, H. G. Allore, J. Dziura, D. Esserman, M. B. Carnie, C. Hanson, F. Ko, N. M. Resnick, J. Wiggins, C. Lu, C. Meng, L. Goehring, M. Fagan, R. Correa-de-Araujo, C. Casteel, P. Peduzzi and E. J. Greene (2021). “Effect of a Multifactorial Fall Injury Prevention Intervention on Patient Well-Being: The STRIDE Study.” J Am Geriatr Soc 69(1): 173–179.

Gregg, E. W., P. Sorlie, R. Paulose-Ram, Q. Gu, M. S. Eberhardt, M. Wolz, V. Burt, L. Curtin, M. Engelgau, L. Geiss and -. n. h. a. n. e. survey (2004). “Prevalence of lower-extremity disease in the US adult population >=40 years of age with and without diabetes: 1999-2000 national health and nutrition examination survey.” Diabetes Care 27(7): 1591–1597.

Guertin, P. A. (2012). “Central pattern generator for locomotion: anatomical, physiological, and pathophysiological considerations.” Front Neurol 3: 183.

Hadjistavropoulos, T., K. Delbaere and T. D. Fitzgerald (2011). “Reconceptualizing the role of fear of falling and balance confidence in fall risk.” J Aging Health 23(1): 3–23.

Hahn, E. A., J. L. Beaumont, P. A. Pilkonis, S. F. Garcia, S. Magasi, D. A. DeWalt and D. Cella (2016). “The PROMIS satisfaction with social participation measures demonstrated responsiveness in diverse clinical populations.” J Clin Epidemiol 73: 135–141.

Hahn, E. A., D. A. DeWalt, R. K. Bode, S. F. Garcia, R. F. DeVellis, H. Correia, D. Cella and P. C. Group (2014). “New English and Spanish social health measures will facilitate evaluating health determinants.” Health Psychol 33(5): 490–499.

Hahn, E. A., M. A. Kallen, R. E. Jensen, A. L. Potosky, C. M. Moinpour, M. Ramirez, D. Cella and J. A. Teresi (2016). “Measuring social function in diverse cancer populations: Evaluation of measurement equivalence of the Patient Reported Outcomes Measurement Information System.” Psychol Test Assess Model 58(2): 403–421.

Halvarsson, A., E. Franzén, E. Farén, E. Olsson, L. Oddsson and A. Ståhle (2013). “Long-term effects of new progressive group balance training for elderly people with increased risk of falling - a randomized controlled trial.” Clin Rehabil 27(5): 450–458.

Hanewinckel, R., J. Drenthen, V. J. A. Verlinden, S. K. L. Darweesh, J. N. van der Geest, A. Hofman, P. A. van Doorn and M. A. Ikram (2017). “Polyneuropathy relates to impairment in daily activities, worse gait, and fall-related injuries.” Neurology 89(1): 76–83.

Hannan, M. T., M. M. Gagnon, J. Aneja, R. N. Jones, L. A. Cupples, L. A. Lipsitz, E. J. Samelson, S. G. Leveille and D. P. Kiel (2010). “Optimizing the tracking of falls in studies of older participants: comparison of quarterly telephone recall with monthly falls calendars in the MOBILIZE Boston Study.” Am J Epidemiol 171(9): 1031–1036.

Hardy, S. E., S. Perera, Y. F. Roumani, J. M. Chandler and S. A. Studenski (2007). “Improvement in usual gait speed predicts better survival in older adults.” J Am Geriatr Soc 55(11): 1727–1734.

Hart, D. A. (2021). “Learning From Human Responses to Deconditioning Environments: Improved Understanding of the “Use It or Lose It” Principle.” Front Sports Act Living 3: 685845.

Hicks, C. W., D. Wang, B. G. Windham, K. Matsushita and E. Selvin (2021). “Prevalence of peripheral neuropathy defined by monofilament insensitivity in middle-aged and older adults in two US cohorts.” Sci Rep 11(1): 19159.

Hoffman, E. M., N. P. Staff, J. M. Robb, J. L. St Sauver, P. J. Dyck and C. J. Klein (2015). “Impairments and comorbidities of polyneuropathy revealed by population-based analyses.” Neurology 84(16): 1644–1651.

Hsu, C. L., I. Iloputaife, L. Oddsson, B. Manor and L. Lipsitz (2021). “Six-Month Lower-Leg Sensory Stimulation Augments Neural Network Connectivity Associated With Improved Gait.” Innovation in aging 5(Suppl 1): 960-961.

Ites, K. I., E. J. Anderson, M. L. Cahill, J. A. Kearney, E. C. Post and L. S. Gilchrist (2011). “Balance interventions for diabetic peripheral neuropathy: a systematic review.” J Geriatr Phys Ther 34(3): 109–116.

Kim, D. H., R. J. Glynn, J. Avorn, L. A. Lipsitz, K. Rockwood, A. Pawar and S. Schneeweiss (2019). “Validation of a Claims-Based Frailty Index Against Physical Performance and Adverse Health Outcomes in the Health and Retirement Study.” J Gerontol A Biol Sci Med Sci 74(8): 1271–1276.

Koehler-McNicholas, S. R., L. Danzl, A. Y. Cataldo and L. I. E. Oddsson (2019). “Neuromodulation to improve gait and balance function using a sensory neuroprosthesis in people who report insensate feet - A randomized control cross-over study.” PLoS One 14(4): e0216212.

Komatsu, H., K. Yagasaki, Y. Komatsu, H. Yamauchi, T. Yamauchi, T. Shimokawa and A. Z. Doorenbos (2019). “Falls and Functional Impairments in Breast Cancer Patients with Chemotherapy-Induced Peripheral Neuropathy.” Asia Pac J Oncol Nurs 6(3): 253–260.

Koski, K., H. Luukinen, P. Laippala and S.L. Kivelä (1998). “Risk factors for major injurious falls among the home-dwelling elderly by functional abilities. A prospective population-based study.” Gerontology 44(4): 232–238.

Kroenke, K., R. L. Spitzer and J. B. Williams (2001). “The PHQ-9: validity of a brief depression severity measure.” J Gen Intern Med 16(9): 606–613.

Kruse, R. L., J. W. Lemaster and R. W. Madsen (2010). “Fall and balance outcomes after an intervention to promote leg strength, balance, and walking in people with diabetic peripheral neuropathy: “feet first” randomized controlled trial.” Phys Ther 90(11): 1568–1579.

Kästenbauer, T., S. Sauseng, H. Brath, H. Abrahamian and K. Irsigler (2004). “The value of the Rydel-Seiffer tuning fork as a predictor of diabetic polyneuropathy compared with a neurothesiometer.” Diabet Med 21(6): 563–567.

Lajoie, Y. and S. P. Gallagher (2004). “Predicting falls within the elderly community: comparison of postural sway, reaction time, the Berg balance scale and the Activities-specific Balance Confidence (ABC) scale for comparing fallers and non-fallers.” Arch Gerontol Geriatr 38(1): 11–26.

Lakens, D. (2013). “Calculating and reporting effect sizes to facilitate cumulative science: a practical primer for t-tests and ANOVAs.” Front Psychol 4: 863.

Li, F., P. Harmer, K. J. Fisher, E. McAuley, N. Chaumeton, E. Eckstrom and N. L. Wilson (2005). “Tai Chi and fall reductions in older adults: a randomized controlled trial.” J Gerontol A Biol Sci Med Sci 60(2): 187–194.

Li, L. and B. Manor (2010). “Long term Tai Chi exercise improves physical performance among people with peripheral neuropathy.” Am J Chin Med 38(3): 449–459.

Lipsitz, L. A., E. A. Macklin, T. G. Travison, B. Manor, P. Gagnon, T. Tsai, I. I. Aizpurúa, O. Y. Lo and P. M. Wayne (2019). “A Cluster Randomized Trial of Tai Chi vs Health Education in Subsidized Housing: The MI-WiSH Study.” J Am Geriatr Soc 67(9): 1812–1819.

Lipsitz, L. A., B. Manor, D. Habtemariam, I. Iloputaife, J. Zhou and T. G. Travison (2018). “The pace and prognosis of peripheral sensory loss in advanced age: association with gait speed and falls.” BMC Geriatr 18(1): 274.

Manor, B., M. Lough, M. M. Gagnon, A. Cupples, P. M. Wayne and L. A. Lipsitz (2014). “Functional benefits of tai chi training in senior housing facilities.” J Am Geriatr Soc 62(8): 1484–1489.

Mathias, S., U. S. Nayak and B. Isaacs (1986). “Balance in elderly patients: the “get-up and go” test.” Arch Phys Med Rehabil 67(6): 387–389.

Melzer, I. and L. I. e. Oddsson (2013). “Improving balance control and self-reported lower extremity function in community-dwelling older adults: a randomized control trial.” Clin Rehabil 27(3): 195–206.

Menz, H. B., S. R. Lord, R. St George and R. C. Fitzpatrick (2004). “Walking stability and sensorimotor function in older people with diabetic peripheral neuropathy.” Arch Phys Med Rehabil 85(2): 245–252.

Meyer, P. F., L. I. Oddsson and C. J. De Luca (2004). “Reduced plantar sensitivity alters postural responses to lateral perturbations of balance.” Exp Brain Res 157(4): 526–536.

Meyer, P. F., L. I. Oddsson and C. J. De Luca (2004). “The role of plantar cutaneous sensation in unperturbed stance.” Exp Brain Res 156(4): 505–512.

Montero-Odasso, M., M. Schapira, E. R. Soriano, M. Varela, R. Kaplan, L. A. Camera and L. M. Mayorga (2005). “Gait velocity as a single predictor of adverse events in healthy seniors aged 75 years and older.” J Gerontol A Biol Sci Med Sci 60(10): 1304–1309.

Montero-Odasso, M. M., N. Kamkar, F. Pieruccini-Faria, A. Osman, Y. Sarquis-Adamson, J. Close, D. B. Hogan, S. W. Hunter, R. A. Kenny, L. A. Lipsitz, S. R. Lord, K. M. Madden, M. Petrovic, J. Ryg, M. Speechley, M. Sultana, M. P. Tan, N. van der Velde, J. Verghese, T. Masud and T. F. o. G. G. f. F. i. O. Adults (2021). “Evaluation of Clinical Practice Guidelines on Fall Prevention and Management for Older Adults: A Systematic Review.” JAMA Netw Open 4(12): e2138911.

Moore, J. L., K. Potter, K. Blankshain, S. L. Kaplan, L.C. O’Dwyer and J. E. Sullivan (2018). “A Core Set of Outcome Measures for Adults With Neurologic Conditions Undergoing Rehabilitation: A CLINICAL PRACTICE GUIDELINE.” J Neurol Phys Ther 42(3): 174–220.

Moreland, B., R. Kakara and A. Henry (2020). “Trends in Nonfatal Falls and Fall-Related Injuries Among Adults Aged ≥65 Years -United States, 2012-2018.” MMWR Morb Mortal Wkly Rep 69(27): 875–881.

Morris, S. B. and R. P. DeShon (2002). “Combining effect size estimates in meta-analysis with repeated measures and independent-groups designs.” Psychol Methods 7(1): 105–125.

Morrison, S., R. Simmons, S. R. Colberg, H. K. Parson and A. I. Vinik (2018). “Supervised Balance Training and Wii Fit-Based Exercises Lower Falls Risk in Older Adults With Type 2 Diabetes.” J Am Med Dir Assoc 19(2): 185.e187-185.e113.

Mustapa, A., M. Justine, N. Mohd Mustafah, N. Jamil and H. Manaf (2016). “Postural Control and Gait Performance in the Diabetic Peripheral Neuropathy: A Systematic Review.” Biomed Res Int 2016: 9305025.

Noohi, F., C. Kinnaird, Y. DeDios, I. S. Kofman, S. Wood, J. Bloomberg, A. Mulavara and R. Seidler (2017). “Functional Brain Activation in Response to a Clinical Vestibular Test Correlates with Balance.” Front Syst Neurosci 11: 11.

Oddsson, L. I. E., T. Bisson, H. S. Cohen, L. Jacobs, M. Khoshnoodi, D. Kung, L. A. Lipsitz, B. Manor, P. McCracken, Y. Rumsey, D. M. Wrisley and S. R. Koehler-McNicholas (2020). “The Effects of a Wearable Sensory Prosthesis on Gait and Balance Function After 10 Weeks of Use in Persons With Peripheral Neuropathy and High Fall Risk - The walk2Wellness Trial.” Front Aging Neurosci 12: 592751.

Oddsson, L. I. E., P. Boissy and I. Melzer (2007). “How to improve gait and balance function in elderly individuals—compliance with principles of training.” Eur Rev Aging Phys Act Volume 4(Issue 1): 15–23.

Patsopoulos, N. A. (2011). “A pragmatic view on pragmatic trials.” Dialogues Clin Neurosci 13(2): 217–224.

Paul, S. S., L. Ada and C. G. Canning (2005). “Automaticity of walking – implications for physiotherapy practice.” Physical Therapy Reviews 10(1): 15–23.

Perera, S., S. H. Mody, R. C. Woodman and S. A. Studenski (2006). “Meaningful change and responsiveness in common physical performance measures in older adults.” J Am Geriatr Soc 54(5): 743–749.

Powell, L. E. and A. M. Myers (1995). “The Activities-specific Balance Confidence (ABC) Scale.” J Gerontol A Biol Sci Med Sci 50A(1): M28–34.

Quigley, P. A., T. Bulat, B. Schulz, Y. Friedman, S. Hart-Hughes, J. K. Richardson and S. Barnett (2014). “Exercise interventions, gait, and balance in older subjects with distal symmetric polyneuropathy: a three-group randomized clinical trial.” Am J Phys Med Rehabil 93(1): 1–12; quiz 13-16.

Reeves, N. D., G. Orlando and S. J. Brown (2021). “Sensory-Motor Mechanisms Increasing Falls Risk in Diabetic Peripheral Neuropathy.” Medicina (Kaunas) 57(5).

Richardson, J. K. and E. A. Hurvitz (1995). “Peripheral neuropathy: a true risk factor for falls.” J Gerontol A Biol Sci Med Sci 50(4): M211–215.

Richardson, J. K., D. Sandman and S. Vela (2001). “A focused exercise regimen improves clinical measures of balance in patients with peripheral neuropathy.” Arch Phys Med Rehabil 82(2): 205–209.

Riskowski, J. L., L. Quach, B. Manor, H. B. Menz, L. A. Lipsitz and M. T. Hannan (2012). “Idiopathic peripheral neuropathy increases fall risk in a population-based cohort study of older adults.” Journal of Foot and Ankle Research 5(1): P19.

Sheldon, J. H. (1948). “Some aspects of old age.” Lancet 1(6504): 621–624.

Shumway-Cook, A., W. Gruber, M. Baldwin and S. Liao (1997). “The effect of multidimensional exercises on balance, mobility, and fall risk in community-dwelling older adults.” Phys Ther 77(1): 46–57.

Sienko, K. H., R. D. Seidler, W. J. Carender, A. D. Goodworth, S. L. Whitney and R. J. Peterka (2018). “Potential Mechanisms of Sensory Augmentation Systems on Human Balance Control.” Front Neurol 9: 944.

Sluijs, E. M., G. J. Kok and J. van der Zee (1993). “Correlates of exercise compliance in physical therapy.” Phys Ther 73(11): 771–782; discussion 783-776.

Soh, S. L., C. W. Tan, J. I. Thomas, G. Tan, T. Xu, Y. L. Ng and J. Lane (2021). “Falls efficacy: Extending the understanding of self-efficacy in older adults towards managing falls.” J Frailty Sarcopenia Falls 6(3): 131–138.

Srinivasan, S. S., G. Tuckute, J. Zou, S. Gutierrez-Arango, H. Song, R. L. Barry and H. M. Herr (2020). “Agonist-antagonist myoneural interface amputation preserves proprioceptive sensorimotor neurophysiology in lower limbs.” Sci Transl Med 12(573).

STEADI, C.-. (2021). “STEADI—Older Adult Fall Prevention.” July 26, 2021. Retrieved December 10, 2021, 2021, from https://www.cdc.gov/steadi/.

Stevens, J. A. and E. A. Phelan (2013). “Development of STEADI: a fall prevention resource for health care providers.” Health Promot Pract 14(5): 706–714.

Streckmann, F., E. M. Zopf, H. C. Lehmann, K. May, J. Rizza, P. Zimmer, A. Gollhofer, W. Bloch and F. T. Baumann (2014). “Exercise intervention studies in patients with peripheral neuropathy: a systematic review.” Sports Med 44(9): 1289–1304.

Strzalkowski, N. D. J., R. M. Peters, J. T. Inglis and L. R. Bent (2018). “Cutaneous afferent innervation of the human foot sole: what can we learn from single-unit recordings?” J Neurophysiol 120(3): 1233–1246.

Studenski, S., S. Perera, K. Patel, C. Rosano, K. Faulkner, M. Inzitari, J. Brach, J. Chandler, P. Cawthon, E. B. Connor, M. Nevitt, M. Visser, S. Kritchevsky, S. Badinelli, T. Harris, A. B. Newman, J. Cauley, L. Ferrucci and J. Guralnik (2011). “Gait speed and survival in older adults.” JAMA 305(1): 50–58.

Studenski, S., S. Perera, D. Wallace, J. M. Chandler, P. W. Duncan, E. Rooney, M. Fox and J. M. Guralnik (2003). “Physical performance measures in the clinical setting.” J Am Geriatr Soc 51(3): 314–322.

Tinetti, M. E. and C. Kumar (2010). “The patient who falls: “It’s always a trade-off”.” JAMA 303(3): 258–266.

Tofthagen, C., C. Visovsky and D. L. Berry (2012). “Strength and balance training for adults with peripheral neuropathy and high risk of fall: current evidence and implications for future research.” Oncol Nurs Forum 39(5): E416–424.

Uitenbroek, D. G. (1997). “SISA-Binomial.” Retrieved May, 2020, from https://www.quantitativeskills.com/sisa/calculations/bonfer.htm.

van Waart, H., M. M. Stuiver, W. H. van Harten, E. Geleijn, J. M. Kieffer, L. M. Buffart, M. de Maaker-Berkhof, E. Boven, J. Schrama, M. M. Geenen, J. M. Meerum Terwogt, A. van Bochove, V. Lustig, S. M. van den Heiligenberg, C. H. Smorenburg, J. A. Hellendoorn-van Vreeswijk, G. S. Sonke and N. K. Aaronson (2015). “Effect of Low-Intensity Physical Activity and Moderate-to High-Intensity Physical Exercise During Adjuvant Chemotherapy on Physical Fitness, Fatigue, and Chemotherapy Completion Rates: Results of the PACES Randomized Clinical Trial.” J Clin Oncol 33(17): 1918–1927.

Vaughan, G. M. (2021). “The roles of mechanoreceptors in muscle and skin in human proprioception.” Current Opinion in Physiology 21: 48–56.

Vinik, A. I., P. Camacho, S. Reddy, W. M. Valencia, D. Trence, A. M. Matsumoto and J. E. Morley (2017). “AGING, DIABETES, AND FALLS.” Endocr Pract 23(9): 1117–1139.

Wang, C., R. Goel, Q. Zhang, B. Lepow and B. Najafi (2019). “Daily Use of Bilateral Custom-Made Ankle-Foot Orthoses for Fall Prevention in Older Adults: A Randomized Controlled Trial.” J Am Geriatr Soc 67(8): 1656–1661.

Winters-Stone, K. M., F. Horak, P. G. Jacobs, P. Trubowitz, N. F. Dieckmann, S. Stoyles and S. Faithfull (2017). “Falls, Functioning, and Disability Among Women With Persistent Symptoms of Chemotherapy-Induced Peripheral Neuropathy.” J Clin Oncol 35(23): 2604–2612.

Wolf, B., H. Feys, De Weerdt, J. van der Meer, M. Noom and G. Aufdemkampe (2001). “Effect of a physical therapeutic intervention for balance problems in the elderly: a single-blind, randomized, controlled multicentre trial.” Clin Rehabil 15(6): 624–636.

Wolf, S. L., R. W. Sattin, M. Kutner, M. O’Grady, A. I. Greenspan and R. J. Gregor (2003). “Intense tai chi exercise training and fall occurrences in older, transitionally frail adults: a randomized, controlled trial.” J Am Geriatr Soc 51(12): 1693–1701.

Wrisley, D. M. and N. A. Kumar (2010). “Functional gait assessment: concurrent, discriminative, and predictive validity in community-dwelling older adults.” Phys Ther 90(5): 761–773.

Wrisley, D. M., G. F. Marchetti, D. K. Kuharsky and S. L. Whitney (2004). “Reliability, internal consistency, and validity of data obtained with the functional gait assessment.” Phys Ther 84(10): 906–918.

Wrisley, D. M., G. McLean, J. B. Hill and L. I. E. Oddsson (2021). “Long-Term Use of a Sensory Prosthesis Improves Function in a Patient With Peripheral Neuropathy: A Case Report.” Front Neurol 12: 655963.

Zaiontz, C. (2020). “Real Statistics Resource Pack (Release 6.8).” Release 6.8. from http://www.real-statistics.com/.

